# Prioritized RNA modification enzymes as risk genes for bipolar I disorder and schizophrenia

**DOI:** 10.64898/2026.01.26.26344826

**Authors:** Bingwu Li, Dan Ohtan Wang

## Abstract

Enzyme-mediated RNA modifications provide a versatile and dynamic regulatory layer in brain development and neural function, orchestrated by over 100 RNA modification proteins (RMPs). A systematic assessment of contribution by common genetic variants in RMP genes to psychiatric disorders has been lacking. Here we curated 123 human RMPs—corresponding to 31 modification types on 6 RNA biotypes—and tested their genetic associations with 16 disorders from the Psychiatric Genomics Consortium (PGC), using GWAS annotation, gene-based association testing, transcriptome-wide association studies (TWAS), Summary data-based Mendelian randomization (SMR), Bayesian colocalization, and gene-set enrichment analysis. As a set of genes, the RMPs did not show significant disease association; at individual gene levels, candidate RMPs were linked to bipolar I disorder (BIPI) and schizophrenia (SCZ). *NSUN2*, a cytosine-5 methyltransferase, was consistently supported by multiple analyses as a risk gene for BIPI and SCZ. Additional candidates, including *NSUN6*, *TYW5*, *TRMT61A*, *QTRT1*, and *MRM1*, also showed significant gene-level associations. All significant SMR associations were negative (β_SMR_ < 0), consistent with a downregulation pattern observed in patient brains. A cross-gene comparison showed that *NSUN2* and *TYW5* exhibit comparable signal enrichment to established risk genes for BIPI and SCZ, respectively. These results warrant future investigations for the roles of specific RMPs in complex mental disorders.

## Introduction

Psychiatric disorders represent some of the most enigmatic and challenging maladies in modern medicine, disrupting core human functions such as perception, cognition, and emotion. Large-scale genomic studies, including genome-wide association studies (GWAS), copy number variation analyses, and epigenomic profiling, have revolutionized the field of major psychiatric disorders by identifying thousands of heritable genetic variants that underlie altered cellular pathways relevant to neuropsychiatric disorders (1). Psychiatric Genomics Consortium (PGC) has taken a leadership role in uniting the global research community and established the world’s largest collaborative framework that enables large-scale, better-powered genetic analysis of neuropsychiatric disorders (including over 400,000 cases and 1.5 million controls) (2). Aligned with this goal, resources such as the CommonMind Consortium, PsychENCODE, BrainSeq, and BrainSpan have provided large-scale profiles of human brain-specific transcriptomic and epigenomic expression that enable systematic interrogation of the regulatory mechanisms underlying neuropsychiatric disease risks (3–6).

However, translating these broad genetic signals into specific biological mechanisms and clinically or therapeutically meaningful insights into potentially treatable diseases remains a daunting task due to their highly complex genetic architectures. Epigenetic regulation has emerged as one of the most important biological frameworks for understanding neuropsychiatric disorders. A landmark PGC pathway analysis identified histone H3-K4 methylation among the top-enriched pathways across schizophrenia (SCZ), bipolar disorders (BIP), and major depressive disorder (MDD), suggesting epigenetic regulation of gene expression as a key vulnerability in neuropsychiatric disorders (7). Methylation quantitative trait loci (mQTL) have been found to be enriched in GWAS signals for BIP and SCZ (8,9).

Beyond DNA and histone modifications, the epitranscriptome represents an additional regulatory layer that has received comparatively less attention in psychiatric genetics. More than 170 modifications have been identified in RNAs, with approximately 50 detected in human cells (10). These modifications impact the production, life cycle, and function of RNAs across a broad range of cellular processes, including transcription, nuclear export, translation, and degradation (11,12). The dynamics of these modifications are governed by specific protein regulators, particularly enzymes that catalyze the modifying (writers) or de-modifying (erasers) reactions at highly conserved nucleotide positions of specific RNAs. Unlike the RNA modifications themselves, which are technically challenging to profile at scale, the coding genes for their corresponding regulatory enzymes can be systematically interrogated in population-level genomic data. However, what genetic risk components are carried by RMPs in the context of neuropsychiatric disorders remains elusive. RMPs are diverse and have not been defined as a functional gene set in standard pathway databases; their collective contribution to the genetic architecture of psychiatric disorders has not been systematically evaluated.

While RNA modifications are present throughout the body, the brain is particularly enriched with modifications such as N6-methyl-adenosine (m6A) (13,14). In addition, the brain harbors a specialized RNA modification landscape with distinctive topological and abundance patterns, suggesting unique functional requirements and operational mechanisms (13,15,16). A large variety of RNA modifications are known to be expressed in the central nervous system and play essential roles for brain development, functions, injury and post-injury repairs (17–20). Human and animal studies have both shown that dysregulated RNA modification leads to neurodegeneration, cognitive deficits, and mental health disorders, such as depression, Fragile X syndrome, ADHD, and Parkinson’s disease, suggesting the requirement of intact function of epitranscriptomic control for maintaining brain function (21–25). A recent multi-omics study integrating m6A-QTL and GWAS data further identified m6A-associated genes across seven neuropsychiatric disorders, underscoring the regulatory role of RNA modifications in psychiatric genetics (26). Studies of familial cases have provided evidence for the dysregulation of RMPs in patients with neurodevelopmental disorders, notably affecting enzymes responsible for transfer RNA (tRNA) modifications (27–30). Synaptically localized m6A-modified mRNAs are encoded by genes associated with neuropsychiatric disorders such as schizophrenia, autism, mood disorders, and intellectual disability (31). Together, these observations position RNA modifications as a functionally important regulatory layer of gene expression in the brain, with growing evidence linking their dysregulation to neuropsychiatric conditions.

Despite the growing recognition of RMPs in human health, a systematic assessment of the genetic contributions by the RMP genes across the landscape of neuropsychiatric disorders is currently lacking. Existing literature has focused on a limited subset of modifications, such as m6A, while the broader involvement of RNA modifications remains largely uncharted. To fill this gap and explore risk contribution by common human genetic variations in RMP genes to neuropsychiatric disorders, we curated a list of 123 known human RMPs corresponding to 31 RNA modifications on 6 RNA biotypes, a growing catalog of epitranscriptomic regulators.

Toward this goal, we applied a multi-method functional genomic pipeline integrating GWAS, gene-based association tests, transcriptome-wide and splicing-based analyses, Mendelian randomization, and colocalization, followed by transcriptomic validation (5,32–37). Through this integrated approach, we prioritized six candidate RMPs based on specific RMP-disease association scores. Our results prioritize *NSUN2* as a consistently supported candidate across multiple analytical methods. We further identified significant associations for *NSUN6*, *TYW5*, *TRMT61A*, *QTRT1*, and *MRM1*.

## Methods and Materials

### Data sources

1. **Reference panel datasets:** For linkage disequilibrium (LD), Mendelian randomization (MR), and colocalization analyses, European-ancestry samples from the 1000 Genomes Project Phase 3 were used as the LD reference panel (38). For fine-mapping analyses, pre-computed LD matrices derived from UK Biobank data were used (39).
2. **Functional QTL resources:** eQTL and sQTL data were obtained from the BrainMeta project, which comprises brain cortex samples from 2,865 individuals, including 2,443 unrelated individuals of European ancestry with genome-wide SNP data (40).
3. **Genome-wide association summary statistics data** for Alzheimer’s Disease (AD), Attention Deficit Hyperactivity Disorder (ADHD), Alcohol dependence (ALCDEP), Autism Spectrum Disorder (ASD), Anxiety Disorder (ANX), Bipolar I-Disorder (BIPI), Bipolar II-Disorder (BIPII), Cannabis use disorder (CUD), Eating Disorders (ED), Hoarding symptoms (HD), Postpartum depression (PPD), Obsessive-compulsive disorder (OCD), Panic disorder (PD), Post Traumatic Stress Disorder (PTSD), Schizophrenia (SCZ), Tourette syndrome (TS) from Psychiatric Genomics Consortium (PGC) and used in accordance with consortium data-use policies (41–55). We acknowledge the PGC investigators for data generation and sharing. Detailed cohort information (ancestry composition and sample size) is summarized in Supplementary Table 2. To match ancestry between GWAS and eQTL reference panels thus improving the accuracy of TWAS analyses, we restricted GWAS summary statistics to European-ancestry samples for the integrative genomic analysis, except for alcohol dependence, for which only multi-ancestry GWAS summary statistics were available.
4. **Transcriptome/RNA-seq datasets**: Gene and isoform-level expression matrices were obtained from the BrainSeq Consortium (56). For schizophrenia, RNA-seq data from the hippocampus and dorsolateral prefrontal cortex of 551 individuals (286 cases and 265 controls) were used, whereas for bipolar disorder, RNA-seq data from the caudate nucleus comprising 44 cases and 266 controls from the same dataset were analyzed.
5. **RNA-modifying proteins (RMPs) gene set**: A total of 123 human RMPs corresponding to 31 RNA modification types were curated from the MODOMICS database, the RNAME database, and the list reported in Delaunay et al., 2024 (10,12,57).

### Gene-based association testing using MAGMA and hMAGMA

Gene and gene set association analyses were performed using SNP-wise = mean models and competitive gene-set analysis in MAGMA v1.10 (32). The 1000 Genomes Phase 3 EUR subpopulation served as reference data for estimating linkage disequilibrium (LD) (38). For the significance of gene-level associations, correlation was performed using the Benjamini & Hochberg method, with an FDR of 0.05 as a threshold. MAGMA and hMAGMA (Hi-C-coupled MAGMA) differ in how SNPs are annotated to genes. In hMAGMA, we extended SNP annotation by incorporating not only exonic and promoter regions but also distal regulatory elements. The distal regulatory regions were defined using Hi-C chromatin interaction data from adult dorsolateral prefrontal cortex (DLPFC) previously (58).

### GWAS fine-mapping

Polygenic functionally informed fine-mapping (PolyFun) was used to annotate the data with per-SNP heritability estimates derived from a meta-analysis of 15 UK Biobank (UKB) traits (39). Neuropsychiatric disorder risk loci were fine-mapped using SuSiE, with the per-SNP heritabilities incorporated as prior causal probabilities (59). Precomputed UKB-based summary LD information was used as the LD reference. Each fine-mapping locus was defined as ±250 kb from the gene boundaries. The SuSiE model assumed a maximum of two causal variants per locus.

### GCTB–SBayesR–based estimation of gene-level heritability contributions

SNP effect sizes were estimated from GWAS summary statistics using the summary-based Bayesian regression (SBayesR) method implemented in GCTB (v2.5.4) (34). Missing summary statistics were imputed in GCTB using a precomputed LD eigen-decomposition based on the UK Biobank European HapMap3 reference panel. SBayesR was run with unscaled genotypes for 20,000 Markov chain Monte Carlo (MCMC) iterations, with a burn-in of 5,000. Gene-level heritability contributions were derived by summing SNP-level variance explained for all SNPs mapped to each gene. SNP–gene mapping was performed based on GENCODE (v26) gene annotations, considering SNPs located within the annotated gene body only. Intergenic regions were not included, and SNPs overlapping multiple genes were assigned to all corresponding genes. The resulting gene-level values were used for subsequent analyses. This conservative gene-body mapping minimized misassignment of non-coding variants, while intergenic regulatory effects were captured by other pipeline components.

### Summary data-based Mendelian randomization (SMR) analysis

SMR (Version 1.3.1) analysis was employed to examine joint associations between GWAS and eQTL data. The 1000 Genomes Phase 3 EUR subpopulation served as the reference data (37,38). The probe window was set to 500 KB. To ensure reliable Mendelian randomization (MR) analyses, MR was performed only for 10 of the 16 PGC disorders for which allele frequency data were available. SNPs included in MR analyses were filtered based on allele frequency criteria: variants were required to have a minor allele frequency (MAF) > 0.01, and SNPs showing allele frequency differences greater than 0.20 between GWAS and eQTL (or sQTL) datasets were excluded. Bonferroni multiple-testing correction was applied to the SMR P-values. Additionally, a post-filtering step applied the heterogeneity in dependent instruments (HEIDI) test, which distinguishes pleiotropy from linkage. Gene–trait associations based on SMR were defined as those with FDR < 0.05 and HEIDI P > 0.05 (indicating no significant heterogeneity and supporting a shared causal variant between the molecular phenotype and the trait). Additional SMR-based integrative results of QTL and trait GWAS associations were obtained from the COLOCdb database (60).

### FUSION-based TWAS analysis

FUSION requires pre-computed SNP-to-expression prediction weights. As such weights are available for the CMC and YFS cohorts but not for larger eQTL references such as BrainMeta, we used the pre-computed CMC Brain-DLPFC (n = 452) and YFS whole blood (n = 1,264) models, in which the optimal predictive model was selected during training based on cross-validated performance among five statistical models (BLUP, BSLMM, LASSO, Elastic Net, and top SNPs) (36). Multiple testing correction for TWAS results was performed using the Benjamini–Hochberg method, with an FDR threshold at 0.05.

### Sparse Partial Least Squares Discriminant Analysis

To visualize the multidimensional relationship between candidate RMP genes and known risk genes, we applied sparse partial least squares (sPLS) using the mixOmics R package to project candidates into a reference space defined by curated control genes (61). For each disorder (SCZ, BIPI, AD, ADHD), we curated disease-specific positive controls and a shared set of negative controls (TYR, FABP4, ALB, TNNT3, OR5M3). A gene-by-feature matrix was constructed from gene-level association statistics across multiple analytical methods, including MAGMA, H-MAGMA, FUSION (DLPFC and blood), SMR (DLPFC), SMR across brain regions, and SMR across cell types, as well as heritability-based features. This was followed by probabilistic PCA (PPCA) for dimensionality reduction and missing value imputation. The sPLS model (two components, five features per component) was trained on the control genes and used to assign each candidate a continuous distance score (0–1) based on its weighted distance to the positive versus negative centroid. This score—intended as a continuous reference projection rather than a classification output—served as one component of the overall prioritization alongside evidence from multiple independent statistical methods.

### Integrative Prioritization Scoring

To integrate evidence across analytical methods, each RMP gene was assigned a cumulative prioritization score. For each of the gene-level analytical approaches (MAGMA, H-MAGMA, SMR, FUSION), predefined empirical weights, following the scoring framework of a previous report (62), were used to score the evidence. The complete weight scheme is provided in Supplementary Table 13. In parallel, a data-driven score was derived from the sPLS-DA analysis, followed by min-max normalization to a 0–1 scale. The final score for each gene was calculated as the sum of the evidence-based weights and the scaled sPLS-DA distance score. Genes were then stratified into “Tier 1” (total score ≥ 10) and “Tier 2” (total score < 10) based on the distribution of the final scores.

### Two-sample MR

For the Two-sample Mendelian Randomization (MR) analysis, SNPs with correlations to RNA expression or splicing events at P < 10e-6 were selected as instrumental variables. The ieugwasr package’s ld_clump function was used for LD clumping of these eQTLs or sQTLs with a cutoff of correlation r² > 0.2 (63). The R package TwoSampleMR was utilized for the analysis: the mr function was used to perform Mendelian randomization. Complementary methods based on different assumptions were used to identify robust associations, including inverse-variance weighting (IVW), MR-Egger, weighted median, and weighted mode. In this study, we primarily relied on the IVW method as the main reference indicator. The mr_heterogeneity function was applied for heterogeneity statistics, and the mr_pleiotropy_test function was used for horizontal pleiotropy statistics (64).

### Colocalization analysis

The coloc package’s coloc.abf function was employed for Bayesian statistical testing in colocalization analysis. Colocalization between GWAS signals and gene eQTLs was assessed using default parameters within a ±500 kb flanking region around each gene. Posterior probabilities were estimated to support five competing hypotheses: PP.H0: No association with either exposure or outcome. PP.H1: Association with exposure only. PP.H2: Association with outcome only. PP.H3: Association with both exposure and outcome, but with different causal variants. PP.H4: Association with both exposure and outcome, sharing a common causal variant. PP.H4 > 0.6 was considered to be significant, and PP.H4 > 0.8 was considered to be strong (35). geni.plots package’s fig_region and fig_stack_region functions were employed to generate LocusZoom plots. Additional coloc-based integrative results of gene-level QTL and trait GWAS associations were obtained from the COLOCdb database (60).

### Multiple Testing Correction Strategy

Each analytical method applied its own significance threshold as described in the corresponding sections. For MAGMA, H-MAGMA, and FUSION TWAS, gene-level associations were corrected using the Benjamini–Hochberg false discovery rate (FDR < 0.05). For SMR, results were required to pass both FDR < 0.05 and the HEIDI heterogeneity test (P > 0.05) to exclude associations driven by linkage rather than a shared causal variant. This dual-filter strategy was applied consistently for both eQTL- and sQTL-based SMR analyses.

The subsequent integrative scoring and sPLS-DA projection were not hypothesis tests but descriptive or prioritization procedures, and thus did not require additional multiple testing correction. This staged approach, in which cross-method consistency serves as the primary safeguard against false positives, follows the framework of a previous multi-method prioritization study (65).

For exploratory COLOCdb analyses, multiple testing correction was not applied, as the primary objective was to calculate signal proportions and enrichment folds rather than to perform formal significance testing. These results should be interpreted as supporting evidence rather than independent statistical validation.

### Bulk RNA-seq Analysis of Prioritized RMP Genes

Gene- and isoform-level expression estimates were obtained from the BrainSeq Phase 2 dataset, processed based on the hg38 reference genome and GENCODE v25 annotations. Differential expression between cases and controls was assessed using the Wilcoxon rank-sum test on log2-transformed Transcripts Per Million (TPM) values.

In addition, we employed logistic regression models with covariates including age, race, mitochondrial read proportion (MitoRate), ribosomal RNA read proportion (rRNArate), total number of detected genes (totalAssignedGene), and RNA integrity number (RIN) to assess whether expression levels are associated with case–control status (schizophrenia or bipolar disorder). The logistic regression model for the case-control analysis is described by the following equation, where Gene Expression is calculated as Transcripts Per Million (TPM), RIN represents the RNA integrity number, and denotes the random error term.

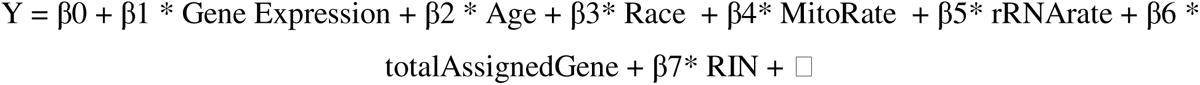

## Results

### Systematic identification of RMP gene associations with PGC disorders

A compilation of 123 human RNA modifying proteins (RMPs) including “writers” and “erasers” corresponding to 31 RNA modifications on 6 RNA biotypes was assembled from the MODOMICS database, the RNAME database, and the gene list reported by Delaunay et al. (10,12,57) (Figure 1a, Supplementary Table 1). To investigate the genetic association of these RMPs across psychiatric conditions, genome-wide association study (GWAS) summary statistics for 16 psychiatric and neurodevelopmental disorders were sourced from the Psychiatric Genomics Consortium (PGC; full list in Supplementary Table 2). As summarized in the descriptive statistics, six disorders: Alzheimer’s Disease (AD), Attention Deficit Hyperactivity Disorder (ADHD), Schizophrenia (SCZ), Bipolar I Disorder (BIPI), Eating Disorders (ED), and Autism Spectrum Disorder (ASD) yielded more than 20 genome-wide significance SNPs across the broad genome-wide analysis (P < 5e-8). The polygenic architecture varied substantially across disorders, with AD displaying a modest polygenic signal and SCZ exhibiting a highly pervasive genome-wide signal (Supplementary Table 2).

**Figure 1:**
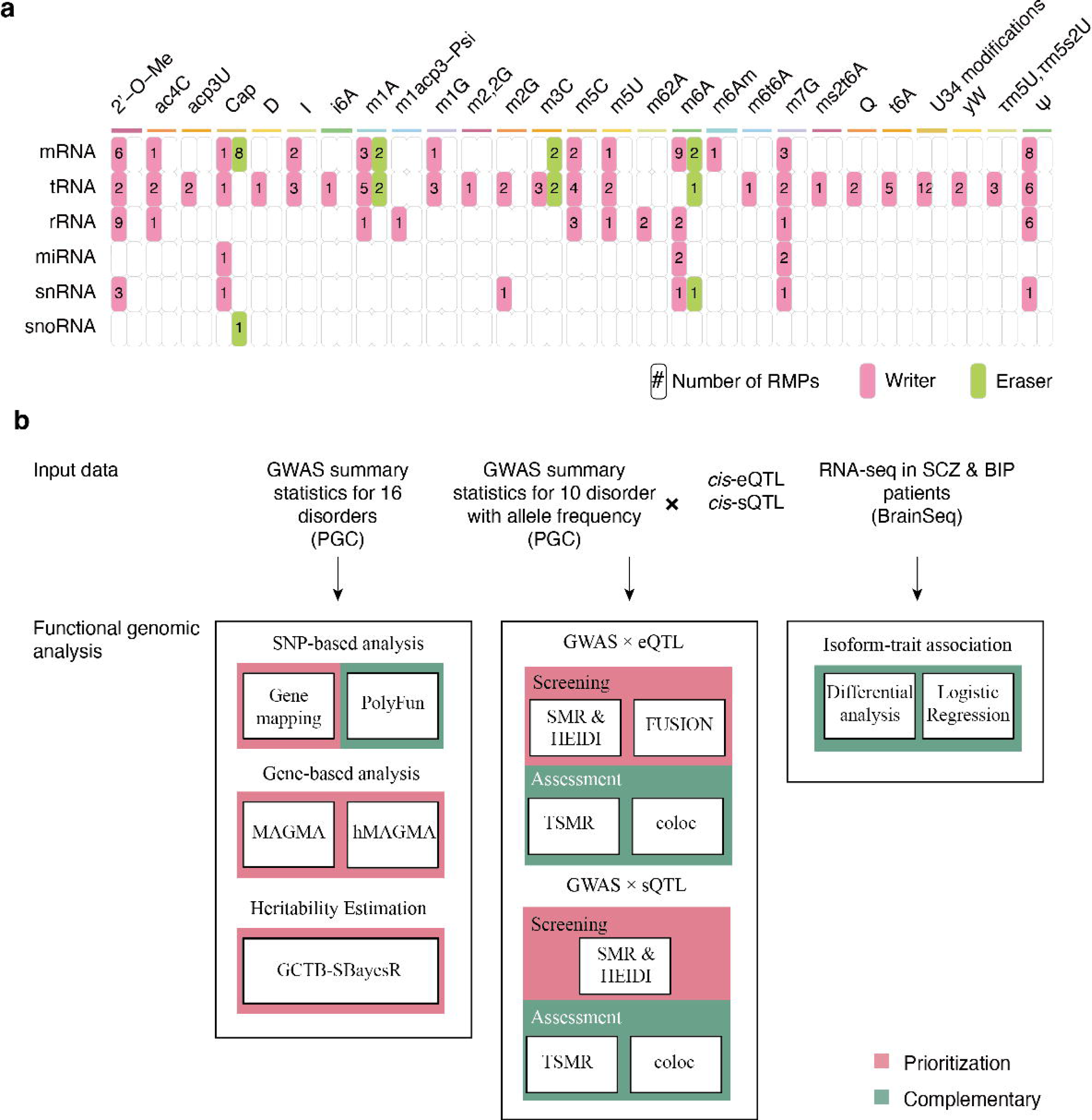
Curated RNA modification proteins (RMPs) and the analytical pipeline. (a) Landscape of the compiled RMPs across 31 distinct RNA modifications (labeled on the top) and various RNA biotypes (labeled on the left), with pre-processed forms categorized under their corresponding RNA biotypes. The numerical values within each cell represent the count of RMPs associated with specific modifications, categorized by their functional roles as “Writers” (pink) or “Erasers” (green). A full RMP list can be found in Supplemental Table 1. (b) Systematic genomic and functional analysis pipeline. The schematic outlines the multi-stage workflow used to identify risk RMP genes for neuropsychiatric disorders.

To systematically identify and prioritize candidate RMP genes associated with PGC disorders, we implemented an integrative pipeline (Figure 1b), beginning with a screening of candidate genes via GWAS signal annotation and gene-based association analyses. Among the 16 PGC disorders, genome-wide significant loci of RMP genes were exclusively found in SCZ (P < 5e-8), specifically involving the loci of *TYW5* and *NSUN6* (Figure S1). Including suggestive signals (P < 1e-6) revealed additional RMP associations in SCZ and BIPI. In SCZ, these included *DCP1A* and *TRMT61A*, whereas in BIPI, these included *NSUN2*, *MRM1*, and *QTRT1* (Figure S1).

We further performed MAGMA and H-MAGMA to conduct gene-based association analyses, which aggregate SNP-level signals across each gene to evaluate its overall genetic association with each disorder. After FDR correction, significant RMP gene associations (FDR < 0.05) were identified in AD, ADHD, BIPI, ED, and SCZ (Figure S2, Supplementary Table 3). The number of significant RMP genes varied markedly across disorders: SCZ exhibited the largest set with 22 RMP genes, followed by BIPI with 8, and AD, ADHD, and ED with 1 each. H-MAGMA yielded similar trends to MAGMA, while identifying a larger number of associated genes (Figure S3, Supplementary Table 4).

We next performed FUSION-based TWAS analysis using expression reference panels from human dorsolateral prefrontal cortex (DLPFC) and blood to identify significant genetically predicted expression–trait associations. In the DLPFC-based analysis, *NSUN2* was associated with both BIPI and SCZ (Figure S4, Supplementary Table 5). Blood-based TWAS detected further associations: *ELP5* in AD and SCZ, *DUS2* and *NSUN2* in BIPI and SCZ (Figure S5, Supplementary Table 6).

To further assess genetically predicted RMP expression and psychiatric disorders, we performed SMR analysis using DLPFC eQTL data. SMR analysis identified significant RMP gene associations with BIPI and SCZ: *MRM2* and *NSUN2* were shared by both disorders, whereas *NAT10*, *TPRKB*, and *TRMT1* showed associations specific to SCZ (Figure S6, Supplementary Table 7). We further extended SMR analyses using eQTL data across multiple brain regions and cell types. *NSUN2* displayed significant associations across all 12 GTEx brain regions, while other RMP genes showed region-specific or disorder-specific associations (Figure S7, Supplementary Table 8). In cell-type-specific analyses, significant SMR associations were identified in BIPI and SCZ across multiple brain cell types (Figure S8, Supplementary Table 9). Extending this framework to the splicing level, SMR analysis based on *cis*-sQTL further identified multiple splicing events of *NSUN2* associated with BIPI, and alternative splicing of *NSUN2*, *THUMPD3*, and *TRMT61B* associated with SCZ (Figure S9, Supplementary Table 10).

Taken together, our multi-method framework identified substantially more RMP associations in BIPI and SCZ compared with other disorders, providing convergent evidence across multiple analytical approaches for RMP involvement in these two conditions (Figure 2a). Fewer RMP associations were detected in AD and ADHD, and none reached significance in the remaining disorders—a pattern that may reflect differences in statistical power across GWAS datasets, genuine biological differences, genetic architecture, or a combination of relevant factors (Figure 2a). GCTB-SBayesR analysis was performed across the four disorders, with genes in the top 5% of heritability contributions considered as supporting evidence (Supplementary Table 11).

**Figure 2:**
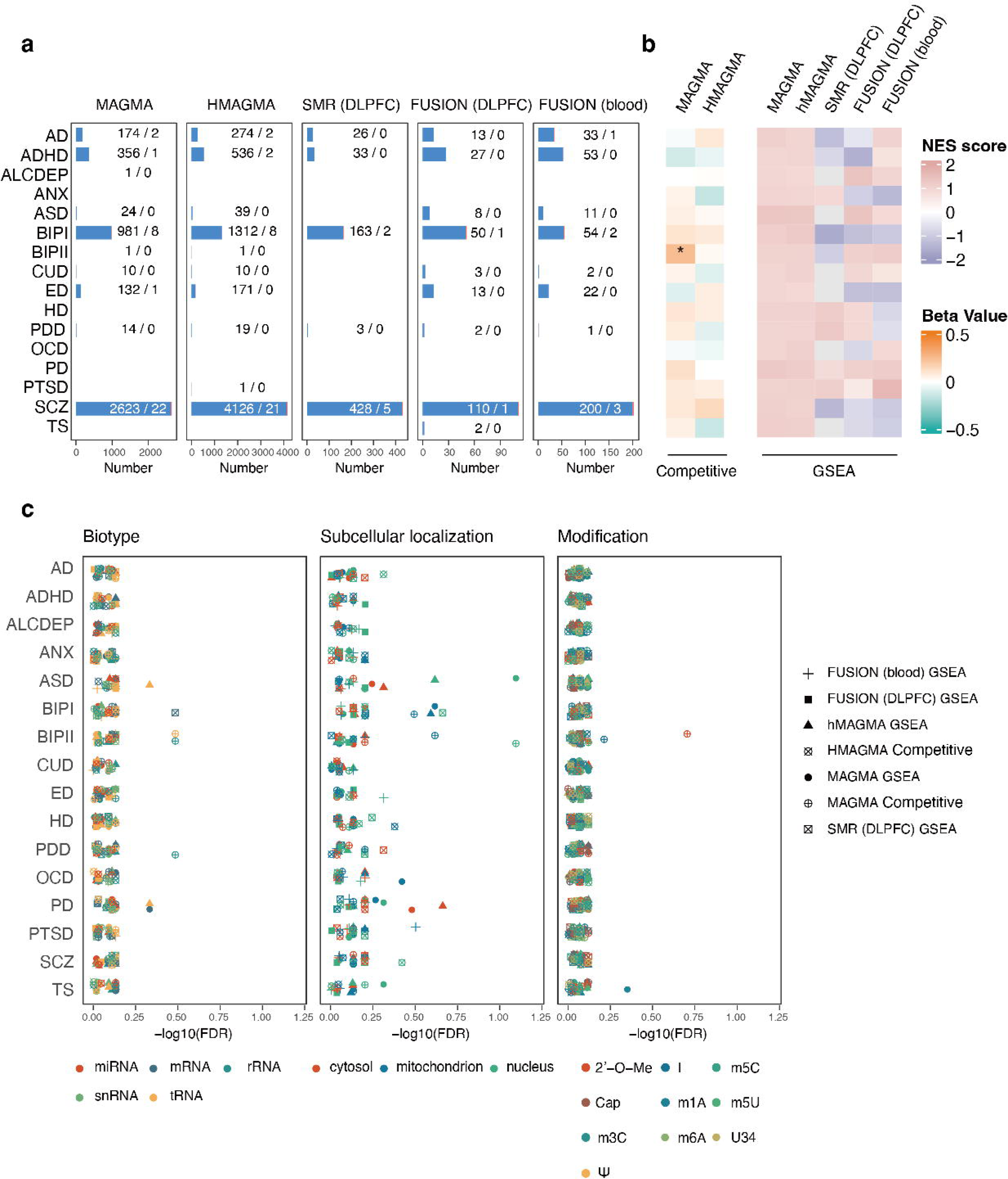
Gene-set enrichment analysis of RMP genes across PGC disorders. (a) Number of significantly associated genes and significantly associated RMP genes (FDR < 0.05) identified across 16 PGC disorders using distinct analytical approaches. (b) RMP gene-set associations across 16 PGC disorders, as determined by competitive gene-set analysis or GSEA. (*: FDR < 0.05). (c) RMP subgene-set associations across 16 PGC disorders, evaluated through competitive gene-set analysis or GSEA.

### Gene set enrichment analysis revealed no significant association between the RMP pathway and psychiatric disorders

To evaluate the aggregate genetic signal of RMP genes, we extended our framework to include gene set enrichment analysis (GSEA) across all four upstream methods (MAGMA, H-MAGMA, SMR, and FUSION), with MAGMA and H-MAGMA further supplemented by competitive gene-set analysis (Supplementary Table 12). Despite the individual RMP gene associations observed in BIPI and SCZ, the RMP gene set as a whole did not show significant enrichment for these disorders in the competitive analyses (FDR > 0.05). Instead, a significant enrichment of association signals was observed in Bipolar II Disorder (BIPII) in the MAGMA competitive gene-set analysis (OR = 1.27, FDR = 0.011; Figure 2b), whereas the self-contained GSEA approaches did not detect significant enrichment for any evaluated disorder (Figure 2b).

To further dissect these signals, we partitioned the RMP gene set into functional sub-gene sets based on their targeted RNA biotypes, subcellular localizations, and modification types (Supplementary Table 12). None of the sub-gene sets reached the FDR-corrected significance threshold (FDR < 0.05), nor showed consistency across analytical methods (Figure 2c). Nucleus-localized RMPs showed nominally suggestive trends in MAGMA-based analyses for ASD and BIPII (FDR = 0.08), but these signals did not survive multiple-testing correction or replicate across methods (Figure 2c).

In summary, the RMP gene set did not exhibit a pervasive association across neuropsychiatric disorders. The lack of broad level gene set enrichment is consistent with the functional heterogeneity of RMPs to modify highly diverse RNA targets for widely ranged downstream signaling processes.

### Seven high-confidence gene–disorder pairs are identified through integrative analysis

In addition to the cumulative evidence gathered from these independent methods, we incorporated sparse partial least squares discriminant (sPLS-DA) modeling analysis, which models gene-level association statistics by projecting them alongside known disease-specific positive and global negative controls. Specifically, we compiled lists of well-established positive control genes from the literature, including *APOE*, *BIN1*, and *TREM2* for AD; *FOXP2* and *ST3GAL3* for ADHD; *CACNA1C*, *ANK3*, *TRANK1*, *SYNE1*, and *TENM4* for BIPI; and *DRD2*, *GRIN2A*, and *CACNA1C* for SCZ. As negative controls, we chose *TYR*, *FABP4*, *ALB*, *TNNT3*, and *OR5M3*. Across all four disorders, the positive and negative control genes showed clear separation in the reference space. Specific RMPs were found in proximity to the positive controls (Figure 3a). For instance, *NSUN2* showed a weighted distance score of 0.53 toward the positive centroid in BIPI, and *TYW5* showed a score of 0.62 toward the positive centroid in SCZ (Figure 3a). RMP candidates did not cluster around positive controls in AD or ADHD. We computed a sPLS-DA distance score based on the candidate’s weighted distances to the centroids of the positive and negative control clusters in the top two components (Figure 3b).

**Figure 3:**
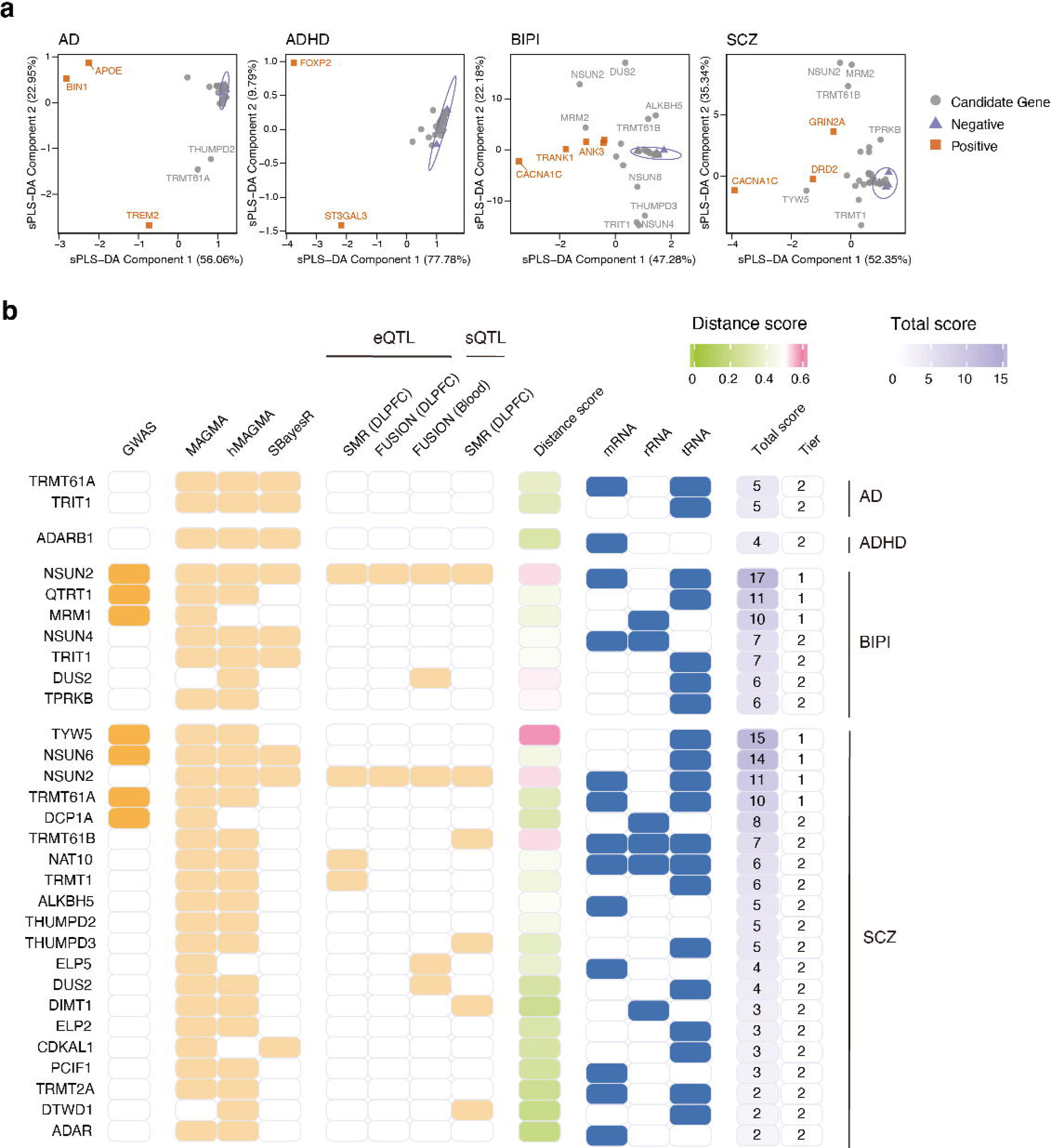
Multi-method prioritization of RMP gene–disorder pairs. (a) sPLS-DA projection of candidate RMP genes (gray) against disease-specific positive (orange) and negative (purple) controls for four psychiatric disorders. Ellipses indicate 95% confidence regions for the control groups. Input features included gene-level association statistics from GWAS-based and QTL-informed analyses (see Methods). (b) Integrative evidence for prioritized RMPs across four disorders: Alzheimer’s Disease (AD), Attention Deficit Hyperactivity Disorder (ADHD), Bipolar I Disorder (BIPI), and Schizophrenia (SCZ), where colored cells display significance in GWAS, gene-based analyses, QTL associations, RNA-seq–based evaluations, and RNA biotype substrates of the prioritized RMPs.

To systematically prioritize RMP gene–disorder pairs across the four disorders (AD, ADHD, BIPI, and SCZ), gene-level evidence from GWAS-based analyses (MAGMA, H-MAGMA, SMR, and FUSION) and the sPLS-DA projection scores were integrated via a weighted sum to yield a total prioritization score. After ranking RMPs by their prioritization scores, the *NSUN2-BIPI* pair ranked the highest, followed by *TYW5*-SCZ, *NSUN6*-SCZ, and *NSUN2*-SCZ (Figure 3b). Identified gene-disease pairs were stratified into Tier 1 (prioritization score >=10) and Tier 2 (Figure 3b).

### Locus-level and cross-trait characterization of RMP candidates

All seven Tier 1 gene–disorder pairs were associated with either BIPI or SCZ, involving six RMPs. With the exception of *MRM1*, which mediates mitochondrial rRNA 2’-O-methylation, the remaining five genes were all involved in tRNA modifications: *NSUN2* and *NSUN6* function as cytosine-5 methyltransferases (m5C), *TYW5* as a Fe² /2-OG-dependent dioxygenase catalyzing wybutosine derivatives, *TRMT1A* as an adenosine-1 methyltransferase (m1A), and *QTRT1* catalyzes queuosine (Q) modification.

We next generated regional association plots for the six disease-associated RMP genes (Figure 4), and performed fine-mapping to assess whether the GWAS signals at these loci converge on the RMP genes. Only *NSUN2-*BIPI locus showed a credible variant mapping within the target gene (rs10454287, PIP > 0.5), whereas fine-mapping signals at other loci were substantially weaker or fell outside the RMP gene region (Figure S10). To evaluate potential causal relationships, we then performed two-sample Mendelian randomization and Bayesian colocalization using eQTL data for the four genes with suitable instrumental variables (*NSUN2*, *NSUN6*, *TYW5*, and *TRMT61A*); *MRM1* and *QTRT1* were excluded from these analyses due to insufficient instrument strength. MR analyses revealed significant associations for *NSUN2* and *NSUN6* in BIPI, and for *NSUN2* and *TYW5* in SCZ (IVW P < 0.05). Colocalization further supported NSUN2 eQTLs correlating to GWAS signals in BIPI (PPH4 = 0.631), whereas no colocalization signals were detected for other gene–disorder pairs (Figure S11 for BIPI, S12 for SCZ). Together, these integrated analyses provided complementary evidence across fine-mapping, MR, and colocalization for the prioritized RMP–disorder pairs.

**Figure 4:**
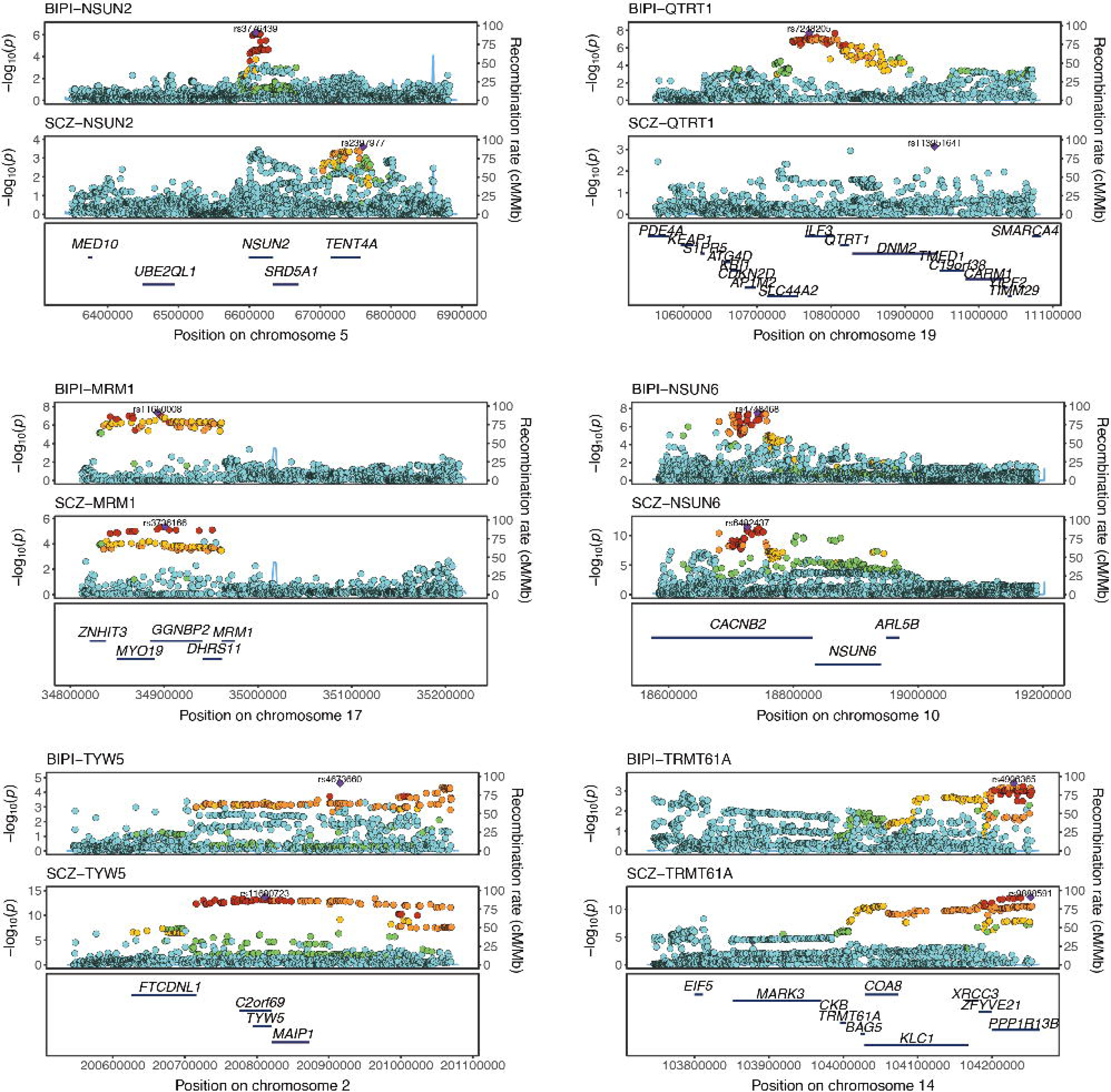
Locus-level association patterns of prioritized RMP–disorder pairs. LocusZoom plots provide a regional view of GWAS association signals for *NSUN2*, *NSUN6*, *TYW5*, *TRMT61A*, *QTRT1*, and *MRM1* in both BIPI and SCZ. For each locus, a ±500 kb genomic region centered on the gene is shown.

We further queried the COLOCdb database to further include molecular quantitative trait loci (xQTLs) in the six prioritized RMP genes in association with BIP and SCZ (60). SMR analysis from COLOCdb yielded highly congruent results with our previous analyses while providing further complementary insights; for instance, *NSUN6* and *TRMT1A* manifested associations with BIP (Figure 5a). Beyond eQTLs and sQTLs, we observed widespread associations with DNA methylation quantitative trait loci (mQTLs)—an epigenetic regulatory layer. Specifically, mQTLs for *NSUN2* were associated with both BIP and SCZ, while mQTLs for *TYW5*, *NSUN6*, and *TRMT1A* were associated with SCZ but not BIP (Figure 5a).

**Figure 5:**
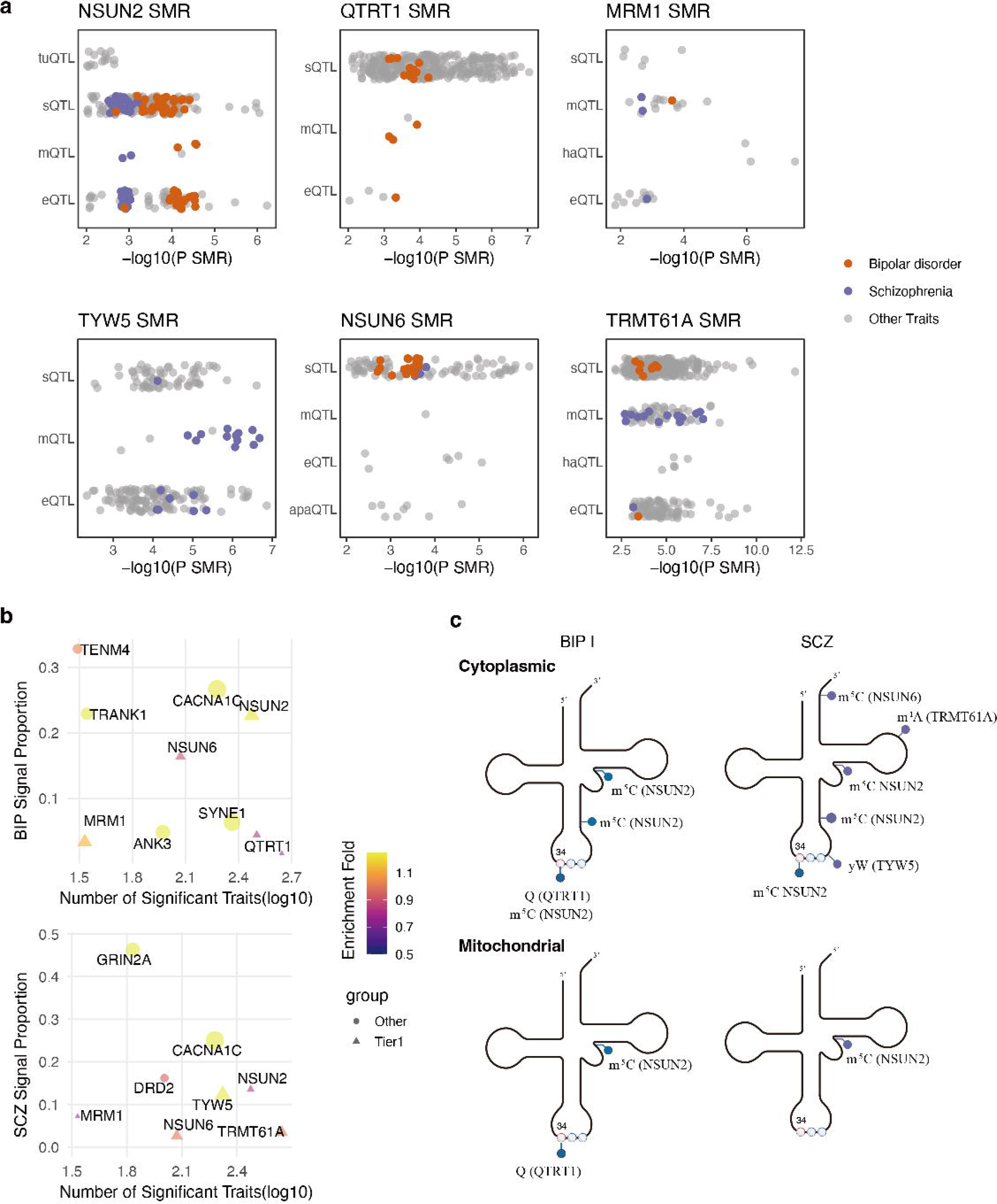
Cross-trait pleiotropic landscape of prioritized RMP genes. (a) SMR analysis results of xQTL–trait associations for the candidate genes. (b) Meta-analysis statistics derived from BIP and SCZ signals in the xQTL–trait SMR results. (c) Modified sites corresponding to tRNA modification-associated genes in BIP and SCZ.

To quantify the pleiotropic effects of these RMP genes and benchmark them against established risk genes, we calculated the signal proportion and enrichment for each gene relative to BIP and SCZ using COLOCdb. For BIP, *NSUN2* exhibited comparable signal proportion and enrichment fold to known BIP risk genes, whereas *NSUN6* showed comparable signal proportion but lower enrichment (Figure 5b). For SCZ, *TYW5* displayed a similar signal proportion and enrichment fold to known SCZ risk genes, whereas *NSUN2* showed comparable signal proportion but lower enrichment (Figure 5b). Following SMR analysis, we further examined colocalization. Among the six RMP genes, *TYW5* and *QTRT1* showed xQTL colocalization with SCZ and BIP, respectively, both of which eluded detection using our SMR–colocalization analyses (Figure S13).

For the five RMPs targeting tRNAs, we mapped their substrate sites onto the secondary structures of cytoplasmic and mitochondrial tRNAs. These include modifications at the anticodon wobble position (position 34: m5C by *NSUN2*, queuosine by *QTRT1*), adjacent to the anticodon (position 37: yW by *TYW5*), and at distal sites including the variable loop (m5C by *NSUN2*), the TψC loop (m1A by *TRMT61A*), and the acceptor arm (m5C by *NSUN6*) (Figure 5c).

### Transcriptomic evidence supports prioritized RMP-disease associations

We next examined the directionality of the RMP–disease associations using SMR effect estimates derived from *cis*-eQTL and GWAS data. Among the six RMP genes, five showed significant SMR associations: *NSUN2*, *TRMT61A*, and *QTRT1* in BIPI, and *NSUN2*, *MRM1*, *TRMT61A*, and *TYW5* in SCZ (Figure 6a). Notably, all significant associations showed negative SMR effect sizes (β_SMR_ < 0), indicating that higher expression of these RMP genes reduces risks for both disorders.

**Figure 6:**
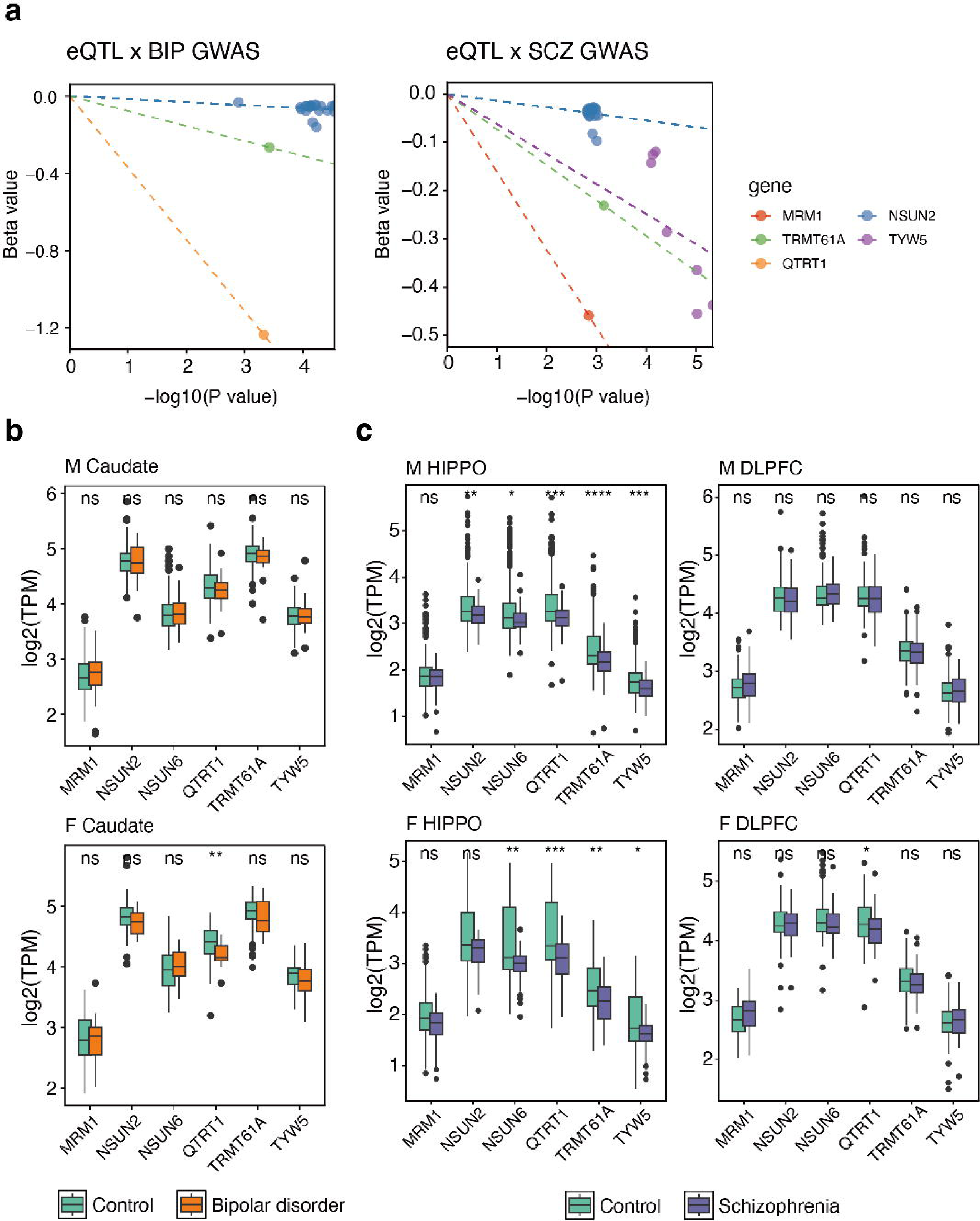
SMR direction and brain expression changes for candidate RMP. (a) eQTL-by-disease interaction analysis results for candidate genes in BIP and SCZ. (b, c) Differential expression analysis of candidate genes in RNA-seq data from BIP (b) and SCZ (c). M: Male; F: Female; HIPPO: Hippocampus; DLPFC: Dorsolateral Prefrontal Cortex.

To assess whether the transcriptional changes are consistent with the genetically predicted variant effects, we examined RNA-seq data sets from the BrainSeq Phase II cohort. Overall, downregulation was observed in patients, consistent with the negative SMR effects. These results were robust to adjustment for relevant technical and demographic covariates in logistic regression models (Figure S14). In BIP, caudate nucleus samples showed significant downregulation of *QTRT1* in female patients (Figure 6b). In SCZ, hippocampal samples showed significant downregulation of *QTRT1*, *TRMT61A*, and *TYW5*, while *NSUN2* downregulation was significant in male patients. In the DLPFC, only *QTRT1* showed significant downregulation, and only in female SCZ patients (Figure 6c).

## Discussion

Various RNA modifications have been identified in the central nervous system (17). In this study, we conducted systematic assessement of RMP genes in association with 16 psychiatric and neurodevelopmental disorders through multi-method functional genomic analysis. Our results show that common variants in human RMP genes carry genetic risks for complex brain disorders. Notably, the strongest evidence supported by multiple analyses results were predominantly identified in BIPI and SCZ. Five out of six identified RMPs encode tRNA modification enzymes: *NSUN2* and *NSUN6* catalyze m5C on tRNAs; *TRMT61A* mediates m1A modification; *QTRT1* catalyzes queuosine modification; and *TYW5* is involved in Wybutosine (yW) formation. Among them, *NSUN6* and *TYW5* have been previously implicated as SCZ risk genes (58), and *QTRT1* has been reported in suggestive GWAS association with bipolar disorder (66), whereas *NSUN2*, *TRMT1A*, and *MRM1* represent novel candidates. While much of the existing literature has focused on the roles of mRNA m6A writers and erasers such as *METTL3* and *FTO*, our results identify a concentration of enzymes specifically targeting tRNA among the high confidence candidates. Given the established roles of these tRNA modifications in translational fidelity and cellular stress responses, their genetic association with BIPI or SCZ warrants further functional investigation.

The identification of *NSUN2* as a shared risk gene for both BIPI and SCZ is particularly significant, as this cytosine-5 methyltransferase is essential for tRNA stability and translational fidelity (67). Biallelic mutations that cause loss of *NSUN2* function have been identified in patients with autosomal recessive intellectual disability, establishing *NSUN2* function as essential for normal brain development (68–70). When *Nsun2* was knocked out from mature neurons in mice, this resulted in selective loss of tRNAGly isodecoders and decreased translational efficiency of glycine-rich synaptic proteins critical for glutamatergic neurotransmission, along with a compensatory increase in cortical glycine levels (71). Given that glycine serves as a co-agonist at the NMDA receptor—a well-established pathophysiological hub in schizophrenia—and that glycine-rich synaptic proteins are essential for glutamatergic signaling, these findings provide a plausible biological link between the genetic risk we observed and the clinical manifestations of BIPI and SCZ (72). Exploratory sQTL-based SMR analysis further suggests that *NSUN2* may confer disease risk partly through alternative splicing and isoform-specific expression, a layer of regulation that remains poorly characterized for this gene.

*NSUN6*, another m5C writer,showed genome-wide association in our analyses. The *NSUN6* function has been strongly linked to neurodevelopment, whose deficiency has been associated with a range of neurodevelopmental disorders, including intellectual disability, autism spectrum disorder, microcephaly, attention-deficit/hyperactivity disorder, and cognitive impairment (73). Nonetheless, studies directly examining the role of *NSUN6* in schizophrenia remain limited. Notably, Wang et al. constructed a regulatory network integrating Hi-C data, QTLs, and activity-by-contact relationships to link noncoding schizophrenia GWAS loci to potential disease genes, in which *NSUN6* was identified as a candidate risk gene (58). More tRNA m5C writers have been linked to SCZ: previous studies reported significant downregulation of *NSUN3* in iPSC-derived cortical interneurons from schizophrenia patients, and *TRDMT1* showed associations with SCZ in prior QTL-based analyses (74).

*TYW5* locus reached genome-wide significance in SCZ, reinforcing the importance of tRNA chemical signatures. Notably, *TYW5* has previously been reported as a schizophrenia risk gene. The earliest large-scale GWAS by Ripke et al. identified a schizophrenia-associated locus on chr2q21.2 that contained *TYW5* gene (75). Subsequently, Li et al. independently confirmed the association of rs796364 and rs281759 with schizophrenia in a Chinese cohort (76). Using CRISPR–Cas9–mediated genome editing, the authors further demonstrated that these two SNPs exert regulatory effects on *TYW5* expression. Functional assays revealed that overexpression of *TYW5* alters neuronal proliferation and differentiation, supporting a role for *TYW5* in neurodevelopment (76). Mechanistically, *TYW5* encodes a tRNA hydroxylase involved in the biosynthesis of wybutosine (yW), a hypermodified guanosine located at position 37 of phenylalanine (Phe)-tRNA. With yW modification playing a critical role for translation fidelity, loss of *TYW5* function may, similar to other members of the wybutosine biosynthetic pathway, impair accurate codon–anticodon pairing of Phe-tRNA. However, the precise mechanisms by which *TYW5* dysregulation contributes to schizophrenia pathogenesis remain unclear. Our findings extend these prior reports by demonstrating that *TYW5* locus reaches genome-wide significance in the latest PGC SCZ GWAS, and by providing complementary evidence at the gene level through MAGMA, SMR, and transcriptomic validation.

*TRMT61A*, encoding an m1A writer that catalyzes the installation of m1A modifications on tRNAs, is located within a schizophrenia-associated locus identified in the earliest large-scale GWAS (75,77). *QTRT1* catalyzes the insertion of queuosine (Q) at the wobble position, a modification essential for translation elongation fidelity. A *Qtrt1* knockout mouse model demonstrated that loss of Q-tRNA modification leads to learning and memory deficits, impaired hippocampal neuronal density, and global imbalances in codon-biased translation elongation (78). Although neither gene has been extensively studied in psychiatric genetics, their associations with SCZ or BIP in our analyses point to m1A and queuosine tRNA modifications as candidates for further functional investigation.

In our SMR analysis, all significant associations showed negative effect sizes (β_SMR_ < 0), indicating that higher expression of these RMP genes is associated with reduced disease risks. This uniform directionality across five RMP genes and two disorders is unlikely to reflect systematic bias, as it is independently supported by the consistent downregulation pattern observed in our BrainSeq RNA-seq validation. It is also consistent with functional evidence from the *Nsun2* knockout model, in which loss of *Nsun2* function leads to synaptic and cognitive deficits (71), suggesting protective roles of these enzymes. It should be noted, however, that the BIP sample (44 cases and 266 controls) in the BrainSeq validation cohort was substantially smaller than the SCZ sample (286 cases and 265 controls), which could have resulted in less statistical power to detect differential expression in BIP.

By identifying promising RMP-disease pairs, we now can prioritize our studies on the disease-relevant pathological context and the function of the specific RMPs, with the goal of translating these genetic signals into clinically relevant biological mechanisms underlying BIP and SCZ. Given that several of the prioritized RMPs are tRNA modification enzymes that play essential roles for tRNA expression and function, which in turn is critical for translational fidelity and tuning protein synthesis, future studies should investigate how their dysregulation affects neuronal protein synthesis —a process known to be essential for synaptic plasticity and cognitive function. tRNA modifications thus emerge as a fundamental biological mechanism to maintain brain health, warranting mechanistic studies in animal models and human brain organoids, to ultimately harness the power of RNA modification regulatory pathways in treating complex brain disorders.

### Limitations of the study

We acknowledge several limitations in our study. First, our genomic analyses were restricted to populations of European ancestry, highlighting the need for future multi-ancestry cohorts to ensure generalizability. Second, statistical power remains a constraint for cross-disorder comparisons; disorders with smaller GWAS sample sizes may yield fewer detectable associations, and the observed difference in signal density between SCZ and other disorders may partly reflect this imbalance. Third, our analyses focused on the genetic association of RMP genes rather than the RNA modifications themselves—modification-level QTLs (e.g., m6A-QTLs) could contribute to disease risk independently of the genes encoding the modifying enzymes, a layer of regulation not captured here. Finally, the heterogeneity of RMP expression and their substrates across brain regions and cell types remains a challenge in connecting this pathway to complex brain diseases. Although we performed exploratory SMR analyses using single-cell and brain-region-specific eQTL data, the limited number of valid instrumental variables at these resolutions constrained the statistical power, and these findings should be interpreted with caution.

## Data availability

This study utilized publicly available datasets. The GWAS summary statistics analyzed in this study are listed in Supplementary Data 1, and detailed information on all other data resources is provided in the Methods section. The shell and R scripts used for data processing, result summarization, and figure generation are available at (https://github.com/lbwfff/PGC_post_GWAS).

## Acknowledgments

The authors would like to thank all lab members in the RNA-MIND Lab and Center for Brain and Health (CBH) at NYUAD for helpful discussions. This research was carried out on the High Performance Computing resources at New York University Abu Dhabi and supported by NYUAD research award to DOW.

