## Supplementary Figure for "Prioritized RNA modification enzymes as risk genes for bipolar I disorder and schizophrenia"

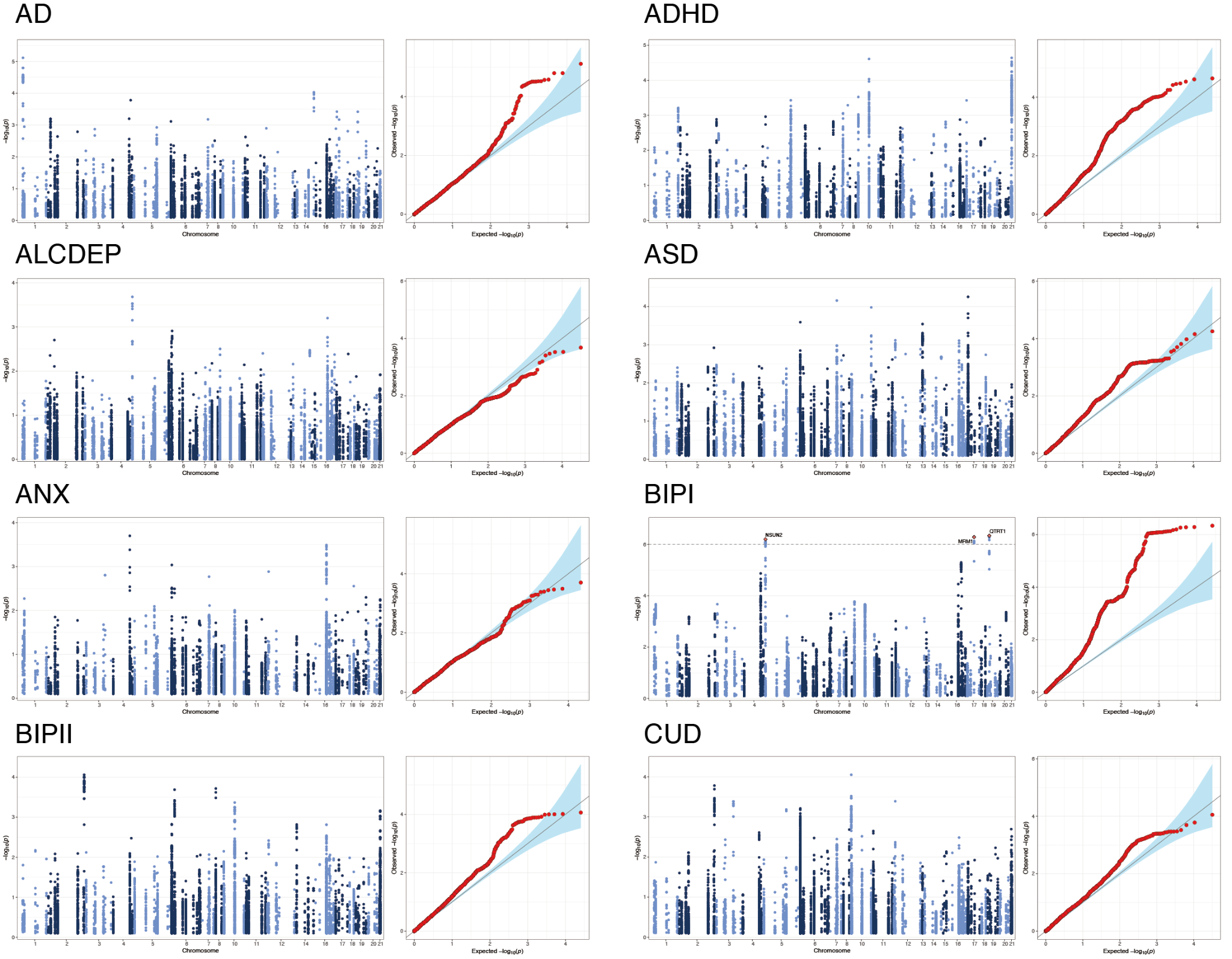

Figure S1: Supplementary figure for Figure 1

Manhattan plots (left) and quantile–quantile (QQ) plots (right) of GWAS summary statistics. (Continued on the next page)

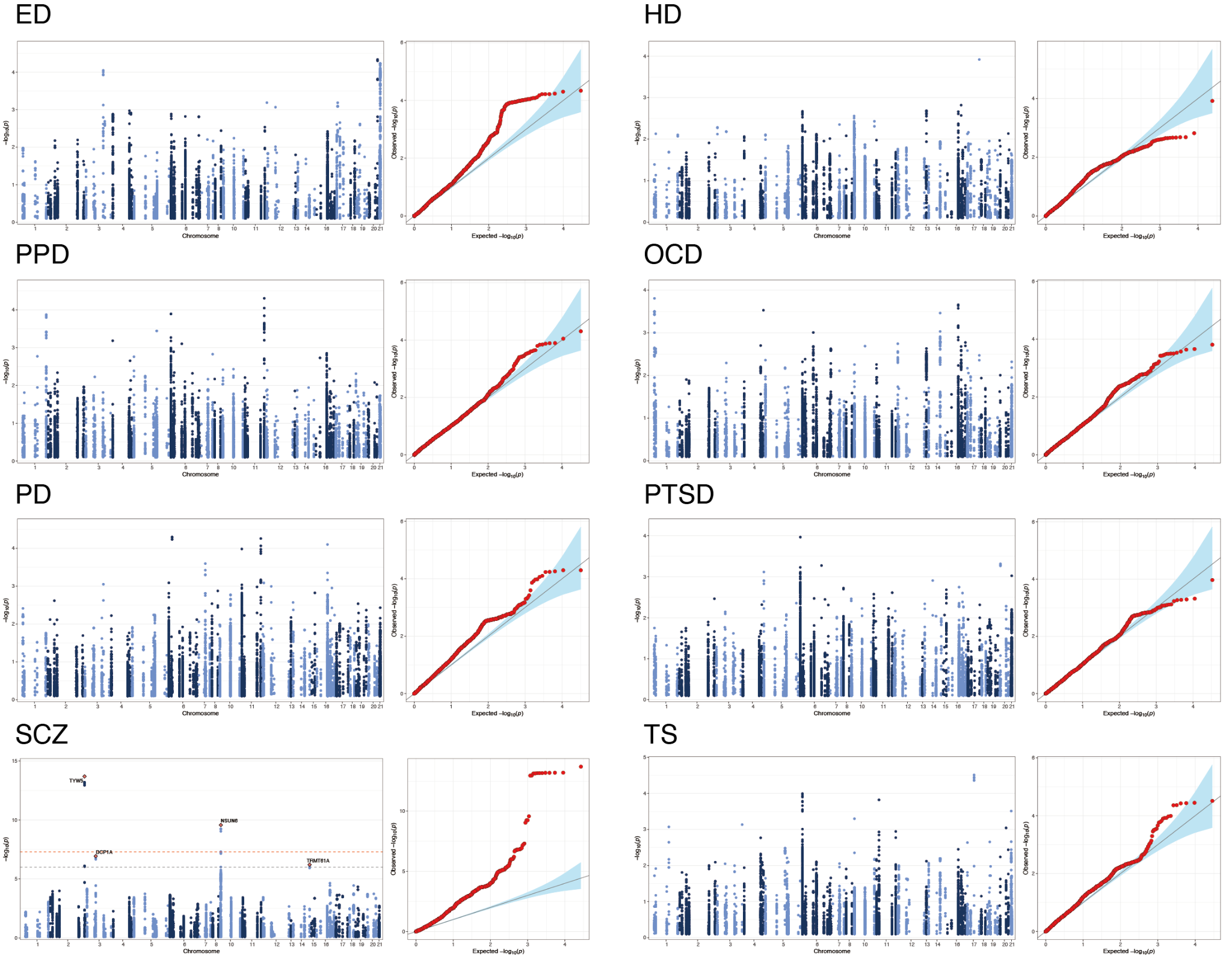

Figure S1 (continued):

Manhattan plots and quantile–quantile (QQ) plots of GWAS summary statistics.

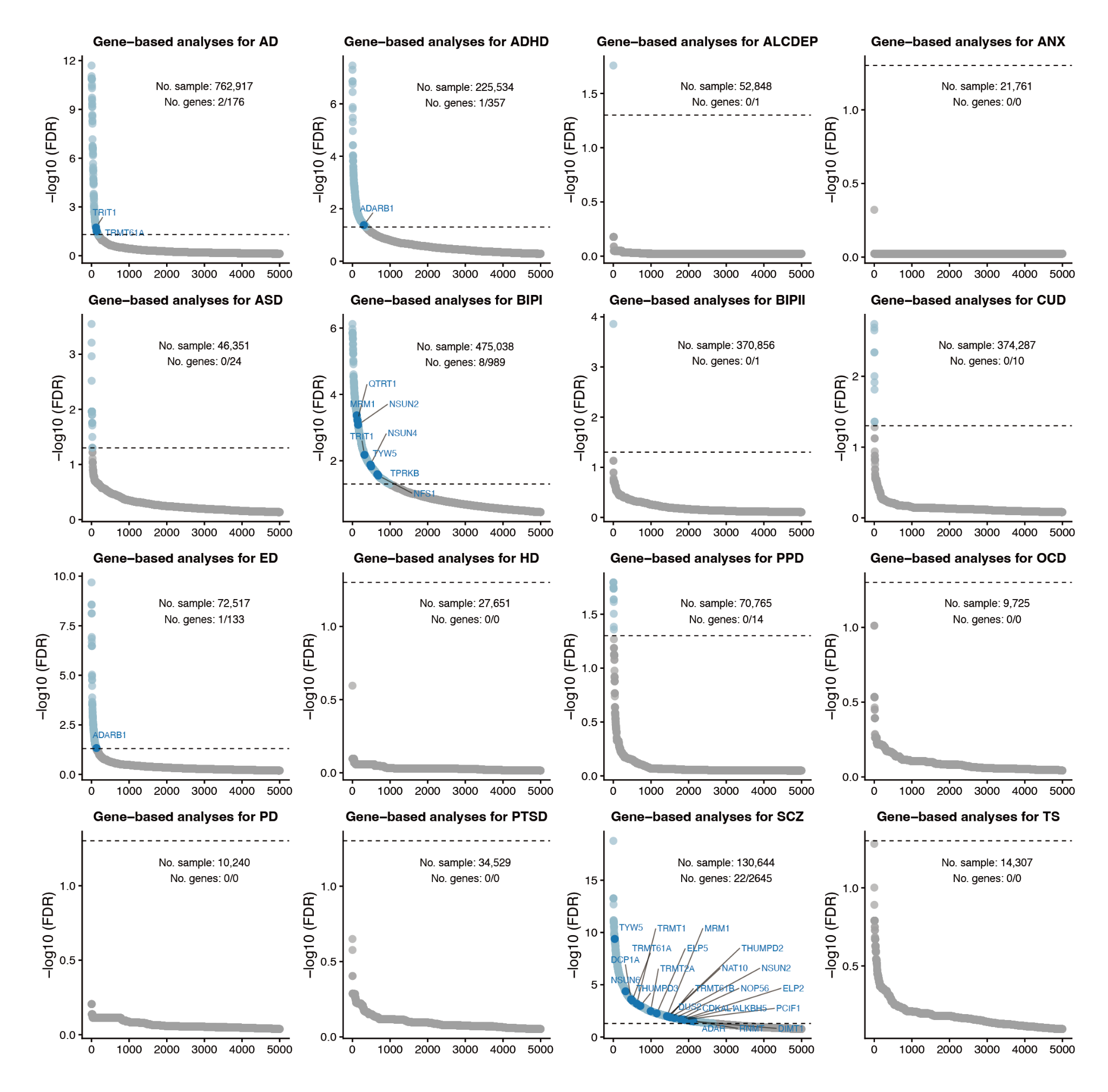

Figure S2: Supplementary figure for Figure 1

Scatter plots illustrating MAGMA gene-based association results for RMP genes. The y-axis represents −log₁₀(FDR), and the x-axis corresponds to genes ranked by −log₁₀(FDR). Annotated dark blue dots represent significantly associated RMR genes; light blue dots represent significantly associated genes (FDR<0.05); gray dots represent non-significant genes (FDR>0.05). Total sample sizes of the GWAS summary statistics (No. sample) and the counts of significantly associated RMP genes and total genes (No. gene) are indicated for each cohort.

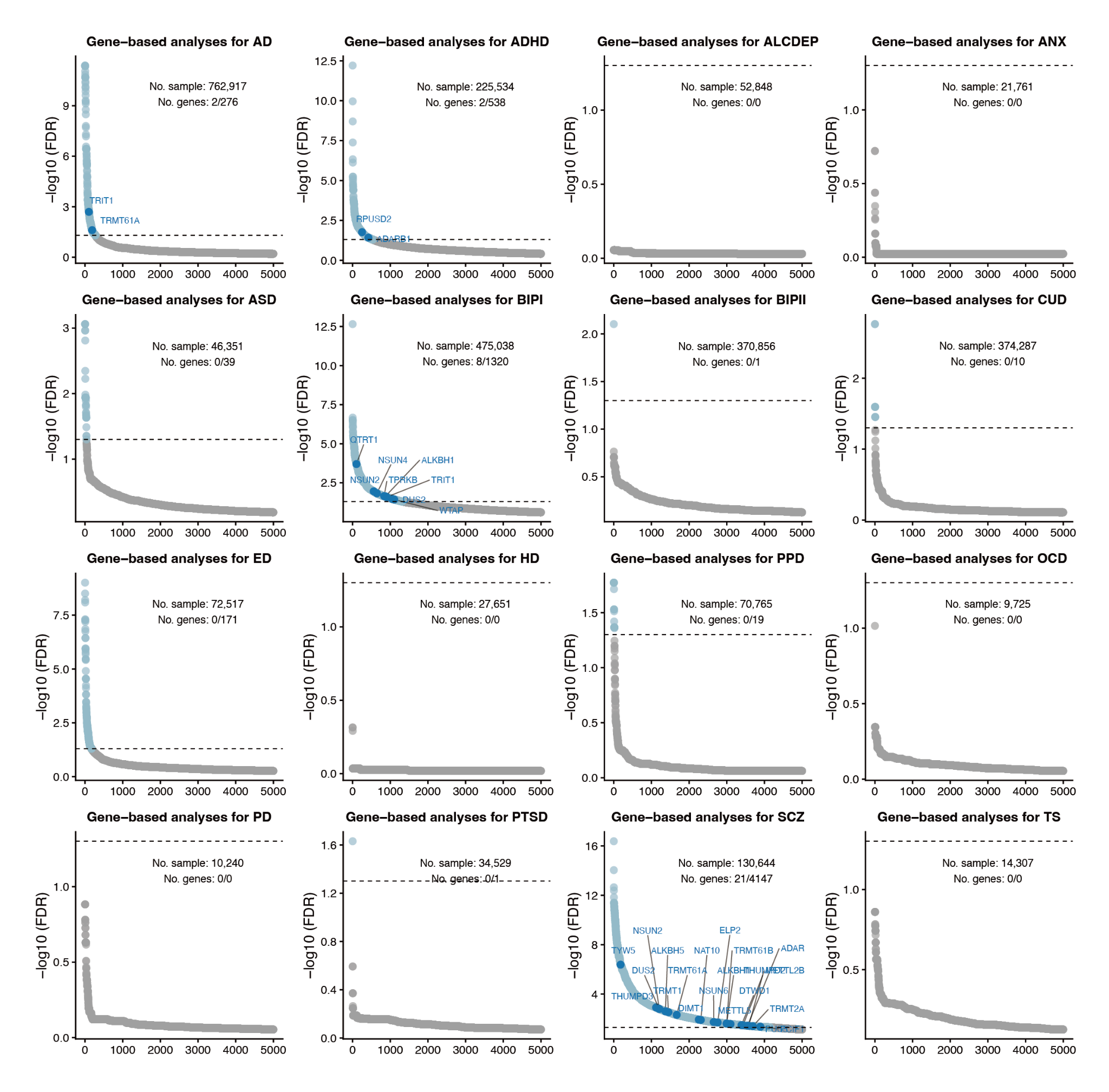

Figure S3: Supplementary figure for Figure 1

Scatter plots illustrating H-MAGMA gene-based association results for RMP genes.

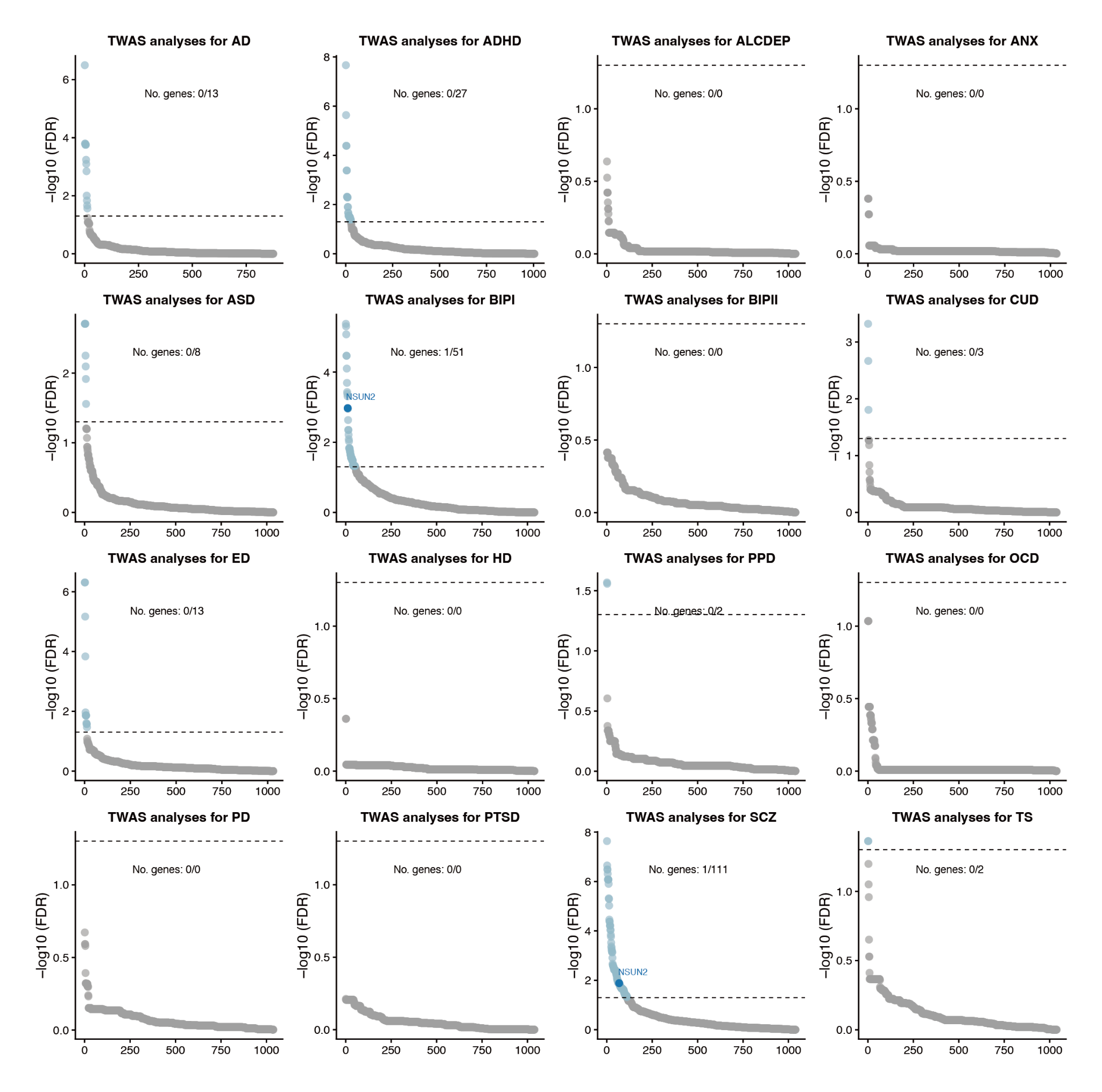

Figure S4: Supplementary figure for Figure 1

Scatter plots illustrating FUSION-based TWAS results based on DLPFC reference panels.

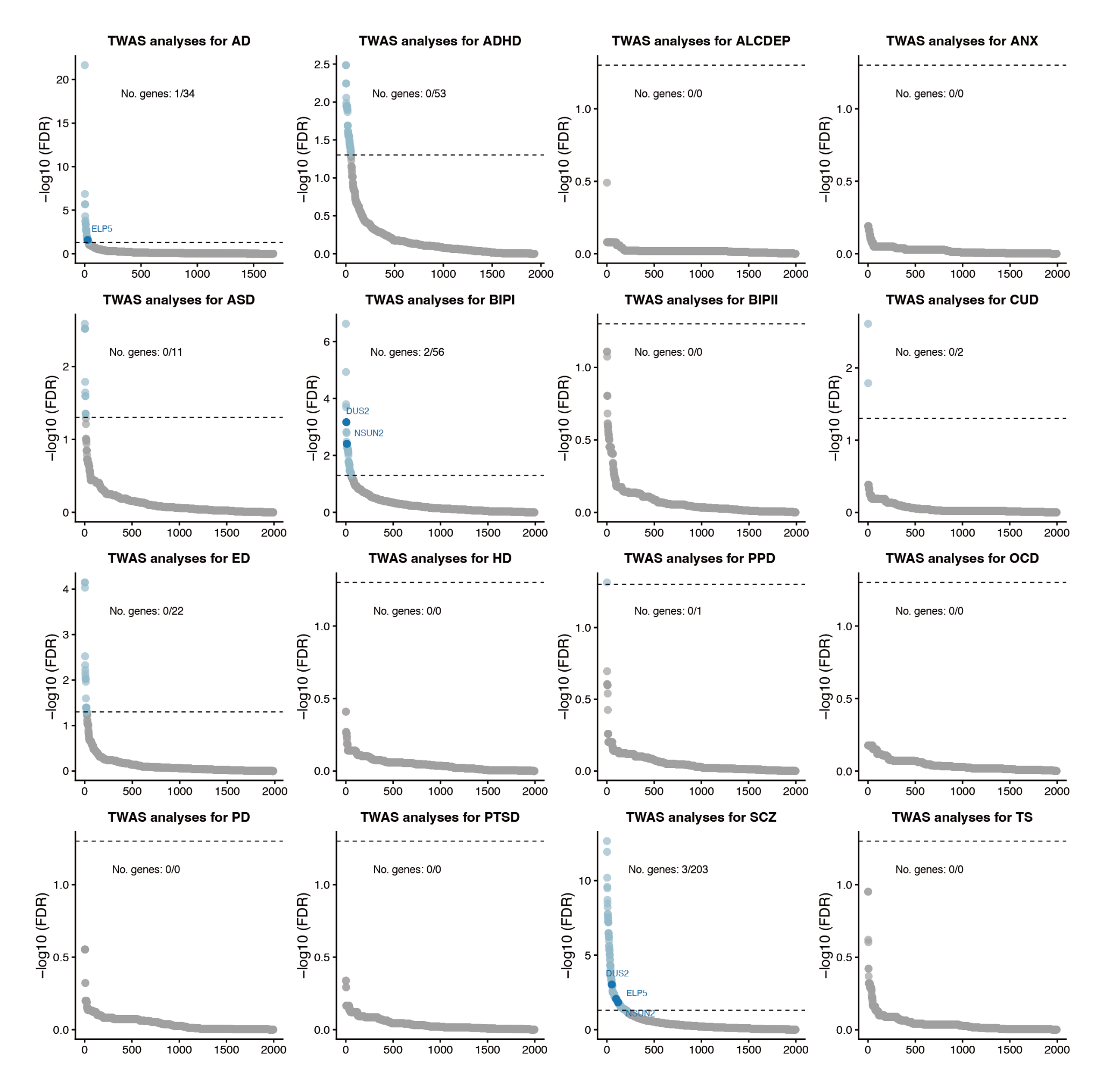

Figure S5: Supplementary figure for Figure 1

Scatter plots illustrating FUSION-based TWAS results based on Blood reference panels.

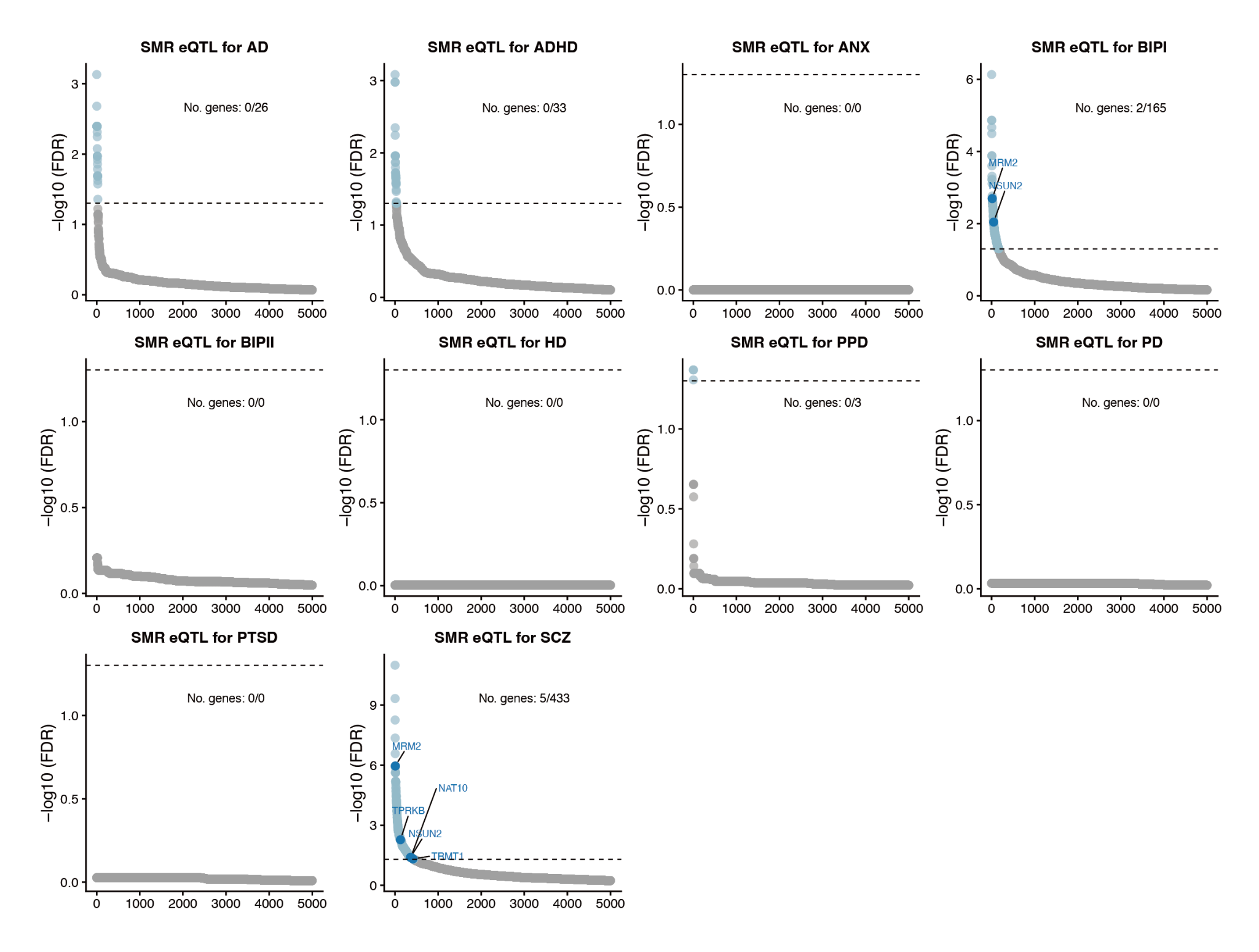

Figure S6: Supplementary figure for Figure 1

Scatter plots illustrating SMR results based on DLPFC eQTL reference panels.

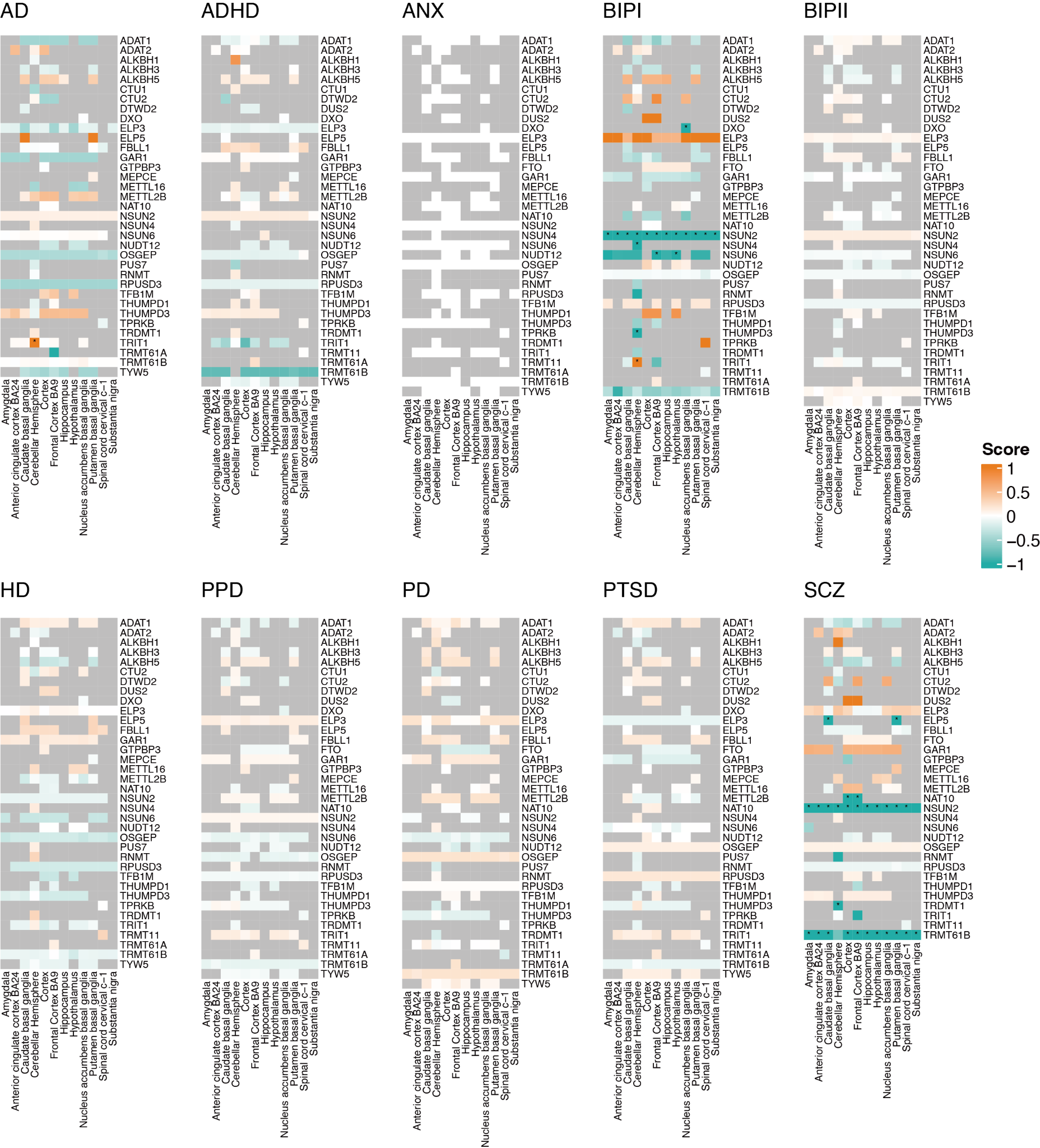

Figure S7: Supplementary figure for Figure 1

SMR analysis results based on eQTLs from different brain regions as references. The score is calculated as -sign(beta)*log10(FDR). Asterisks indicate significant associations with FDR < 0.05.

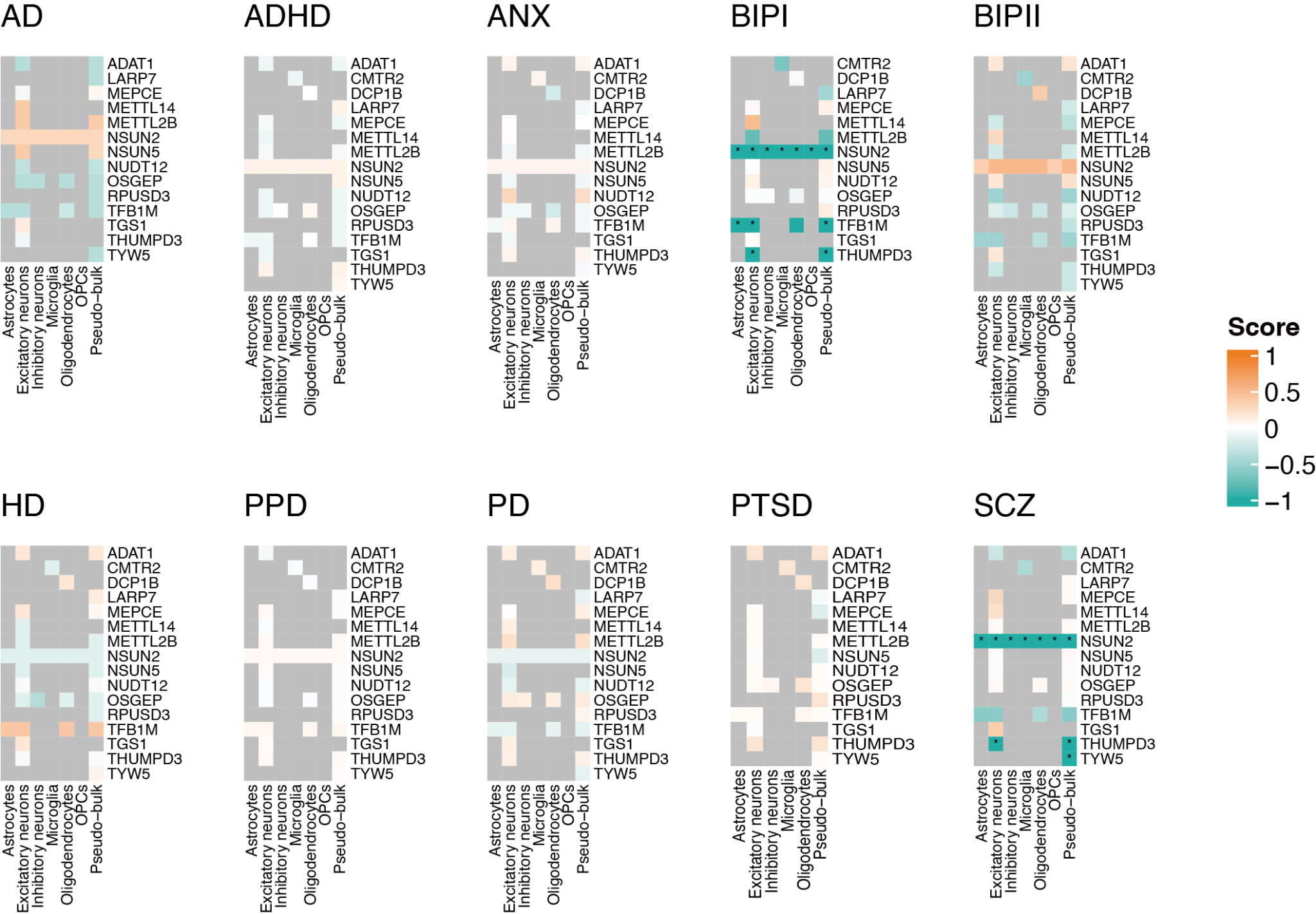

Figure S8: Supplementary figure for Figure 1

SMR analysis results based on eQTLs from different cell types as references. The score is calculated as -sign(beta)*log10(FDR). Asterisks indicate significant associations with FDR < 0.05.

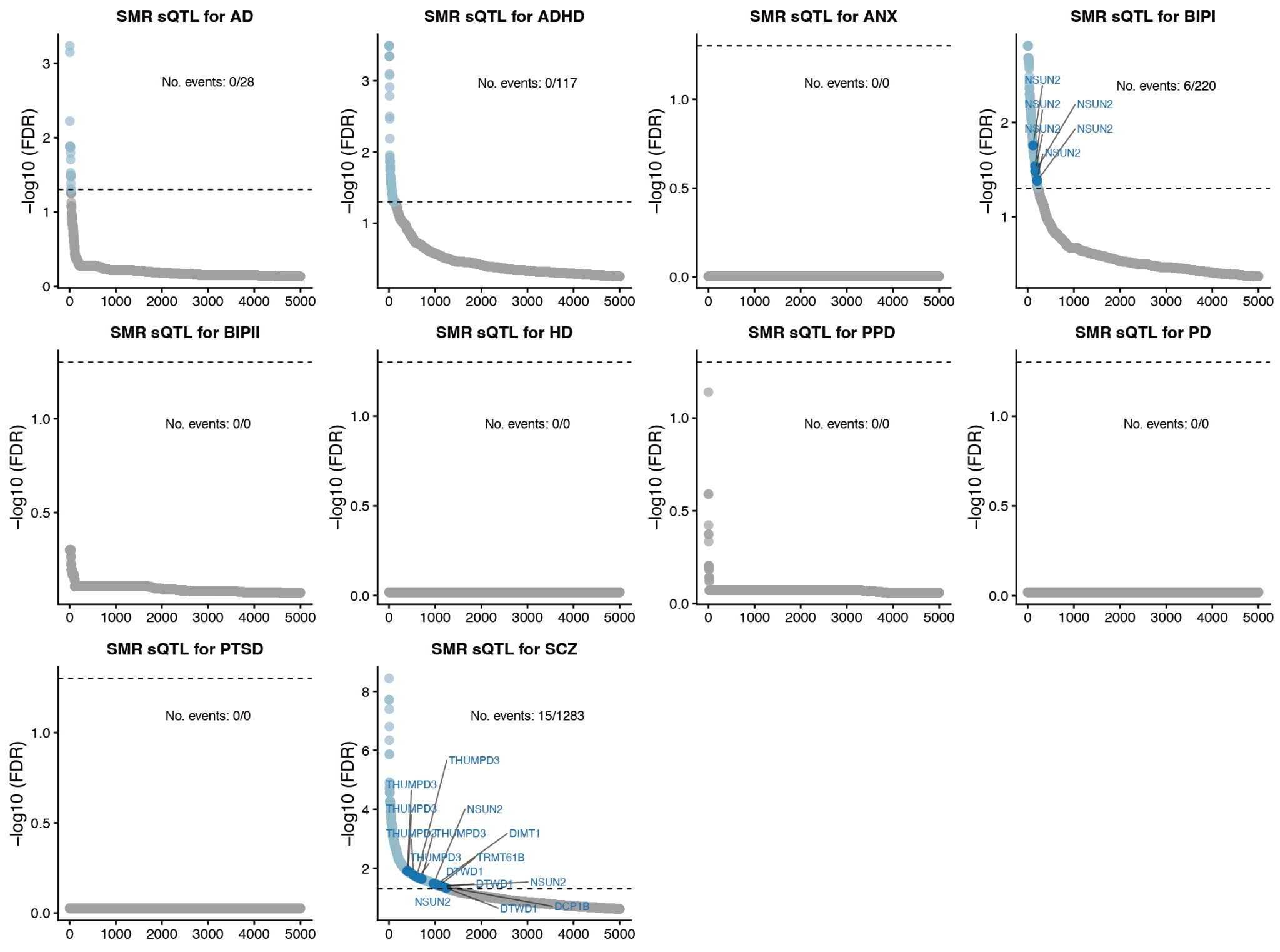

Figure S9: Supplementary figure for Figure 1

Scatter plots illustrating SMR results based on DLPFC sQTL reference panels.

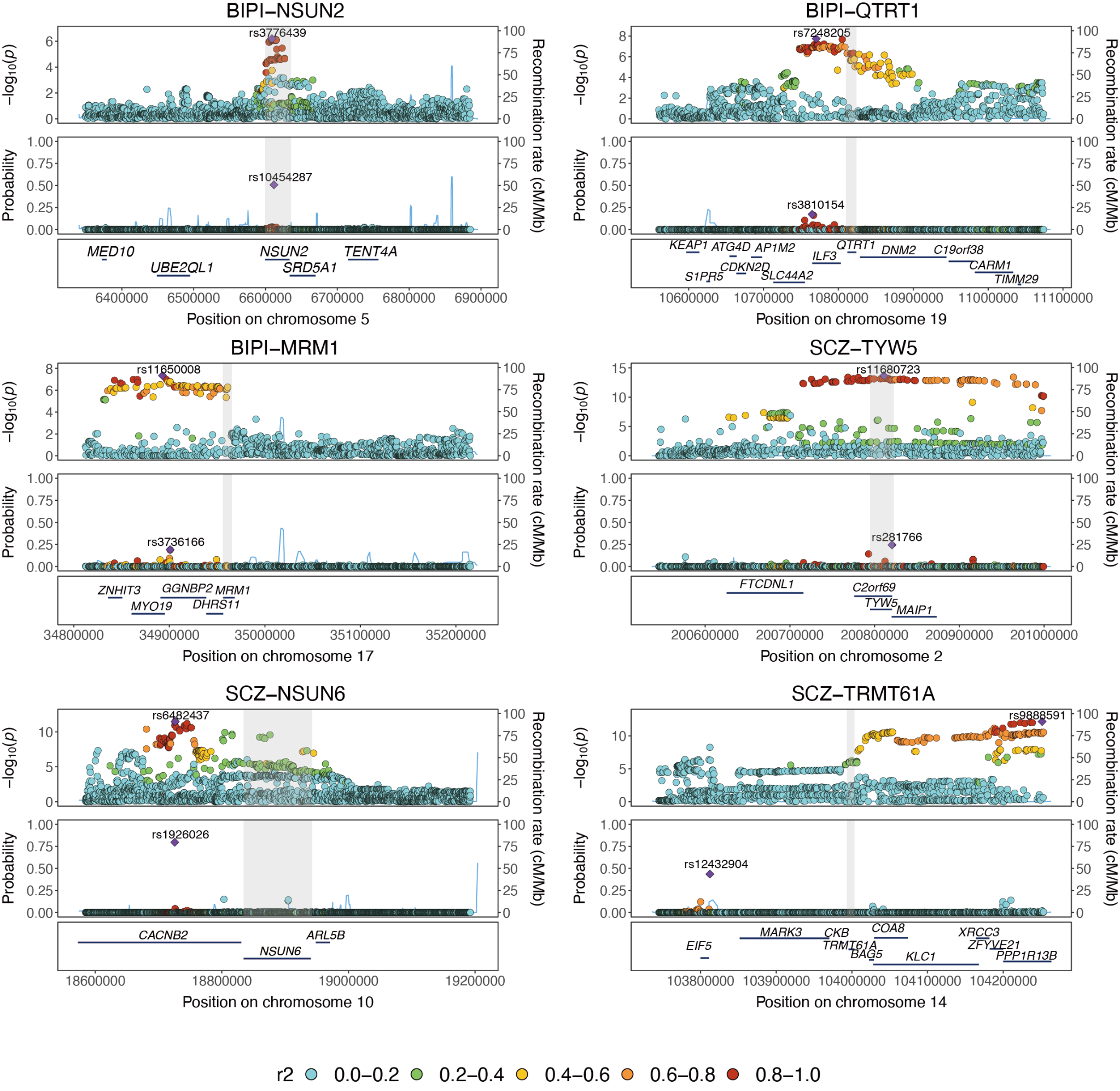

Figure S10: Supplementary figure for Figure 4

Stacked LocusZoom plots provide a regional view of GWAS signals and fine-mapping posterior inclusion probabilities (PIPs) for *NSUN2* (BIPI), *QTRT1* (BIPI), *MRM1* (BIPI), *TYW5* (SCZ), *NSUN6* (SCZ), and *TRMT61A* (SCZ). For each region, a ±500 kb genomic region centered on the gene locus is shown in the LocusZoom plots. Gray rectangles indicate the gene body and promoter regions within the genomic locus.

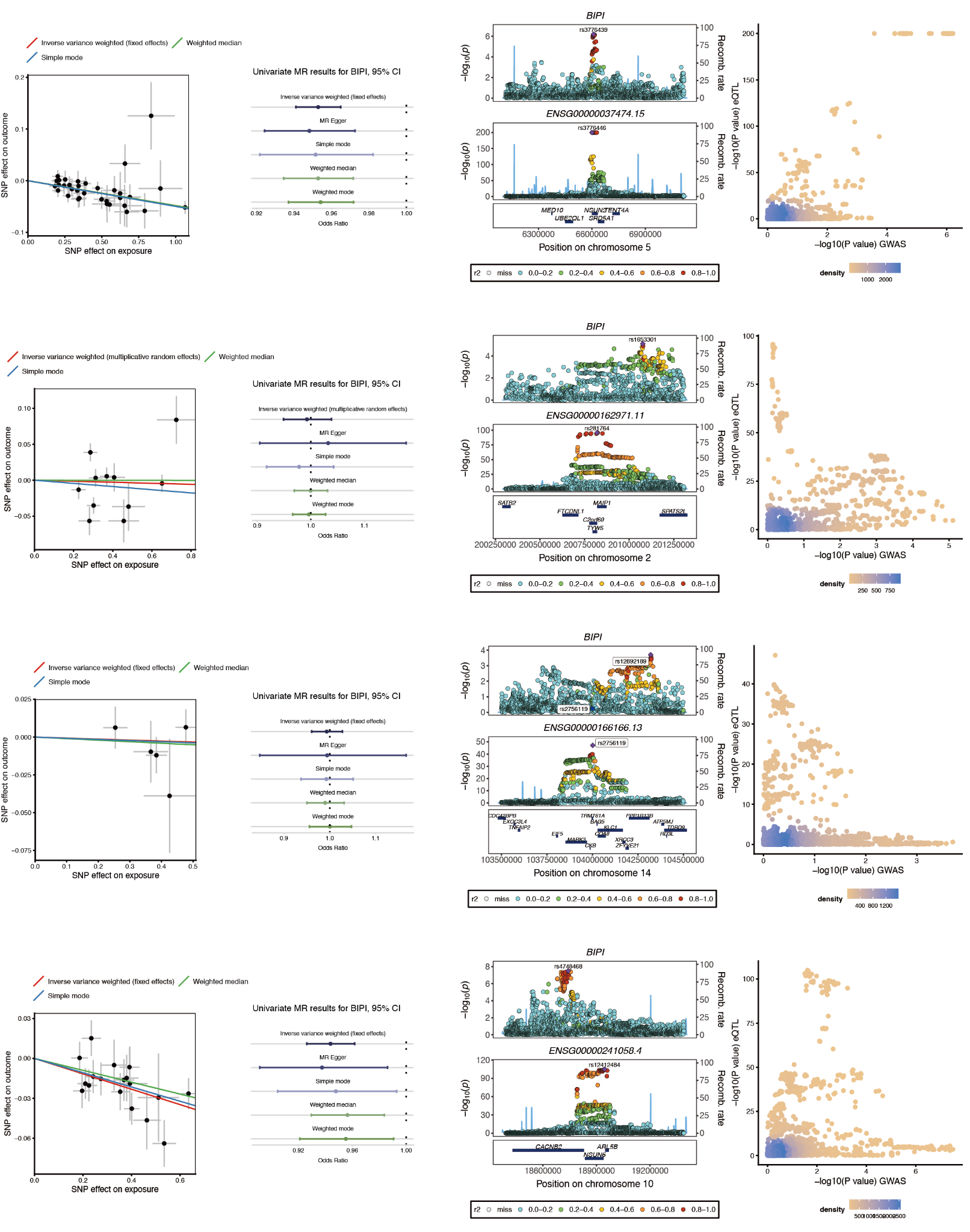

Figure S11: Supplementary figure for Figure 4

Two-sample Mendelian randomization and colocalization analyses for prioritized RMP genes in BIPI. Results are shown for *NSUN2*, *TYW5*, *TRMT61A*, and *NSUN6*. Left panels: scatter plots depicting GWAS and eQTL effect sizes for individual SNPs, with regression lines for each MR method; the slope represents the estimated causal effect. Middle panels: forest plots summarizing the estimated causal effects from different MR methods. Right panels: colocalization LocusZoom plots with corresponding scatter plots of GWAS versus eQTL association P-values.

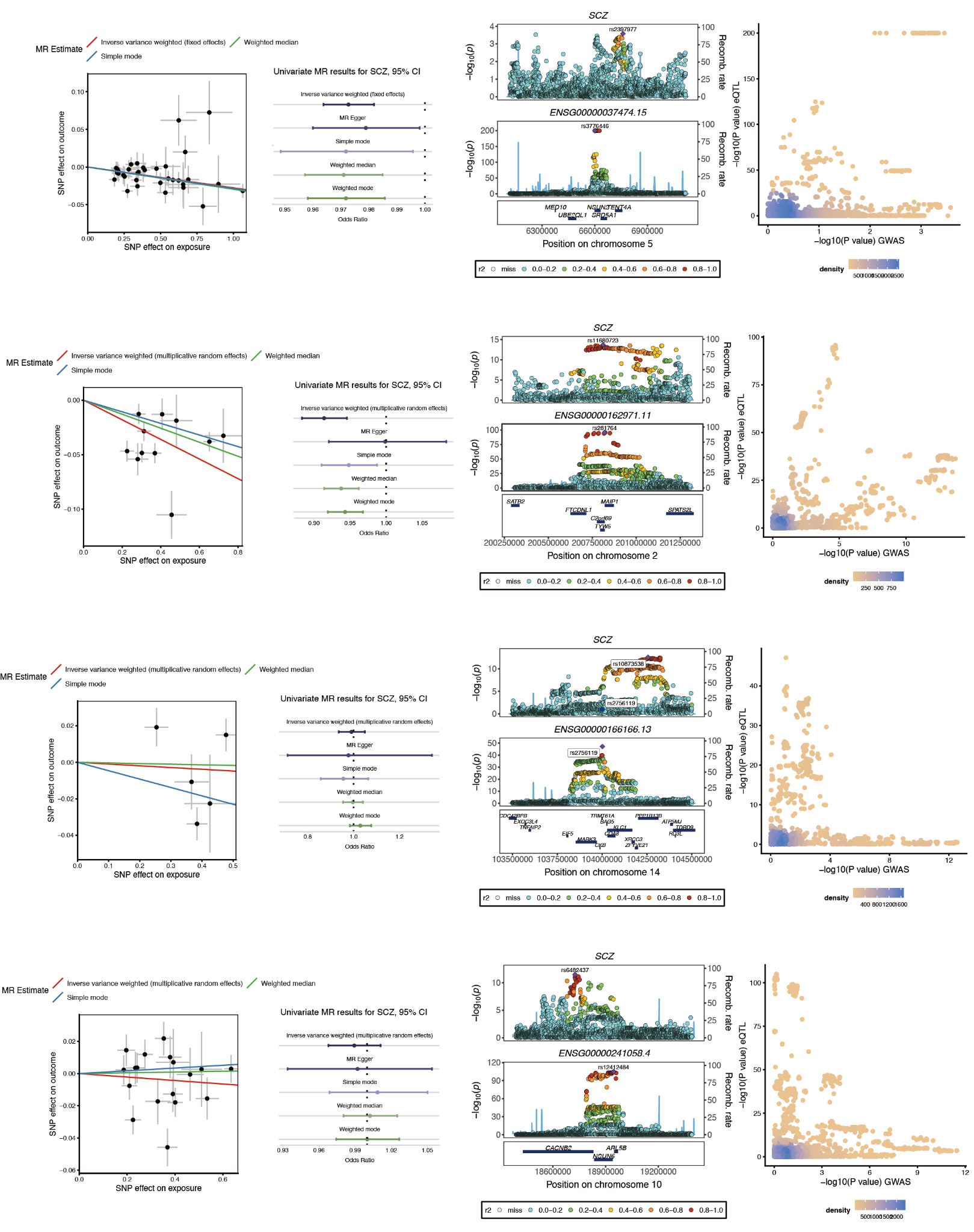

Figure S12: Supplementary figure for Figure 4

Two-sample Mendelian randomization and colocalization analyses for prioritized RMP genes in SCZ.

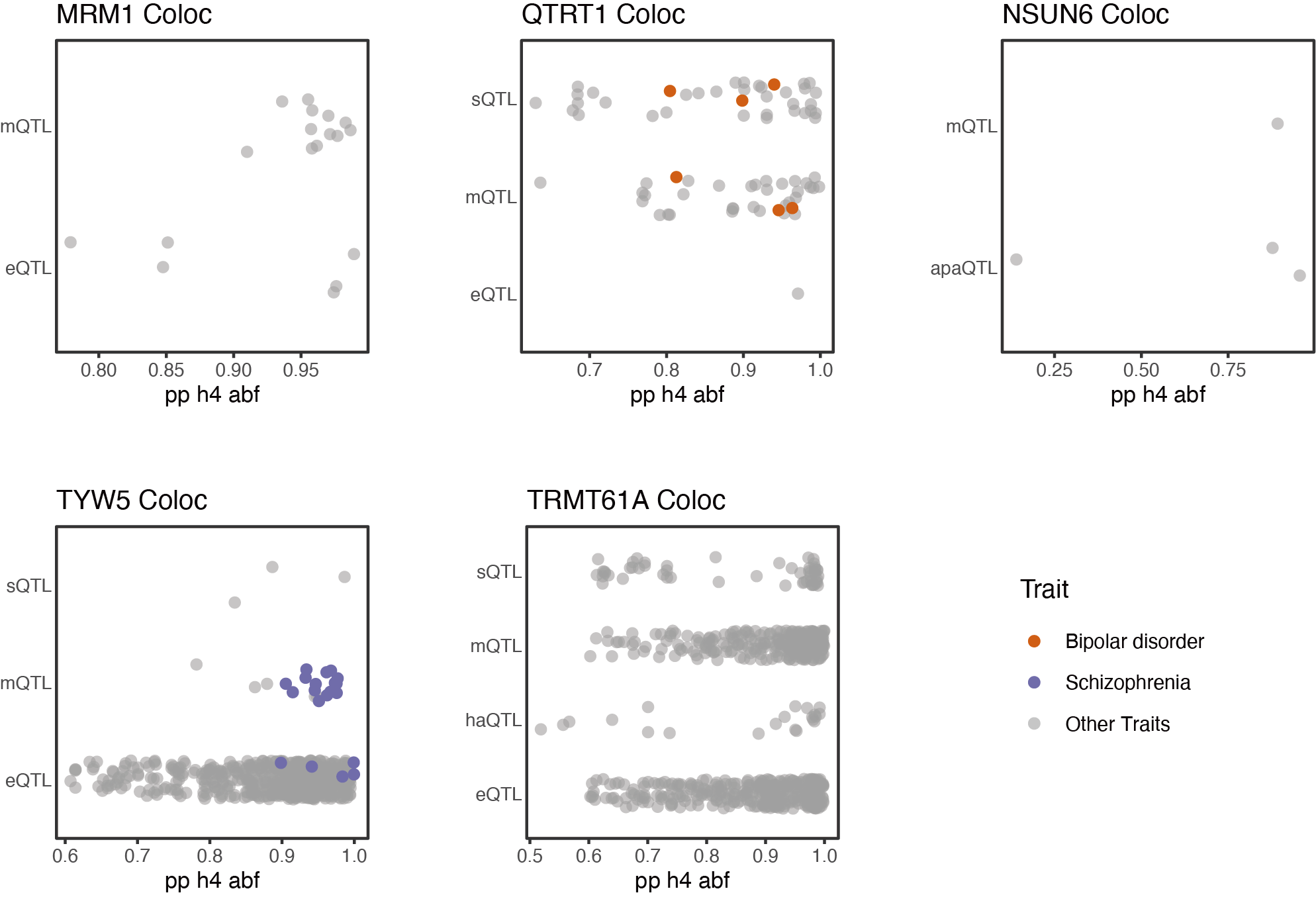

Figure S13: Supplementary figure for Figure 5

Coloc analysis results of xQTL–trait associations for the candidate genes

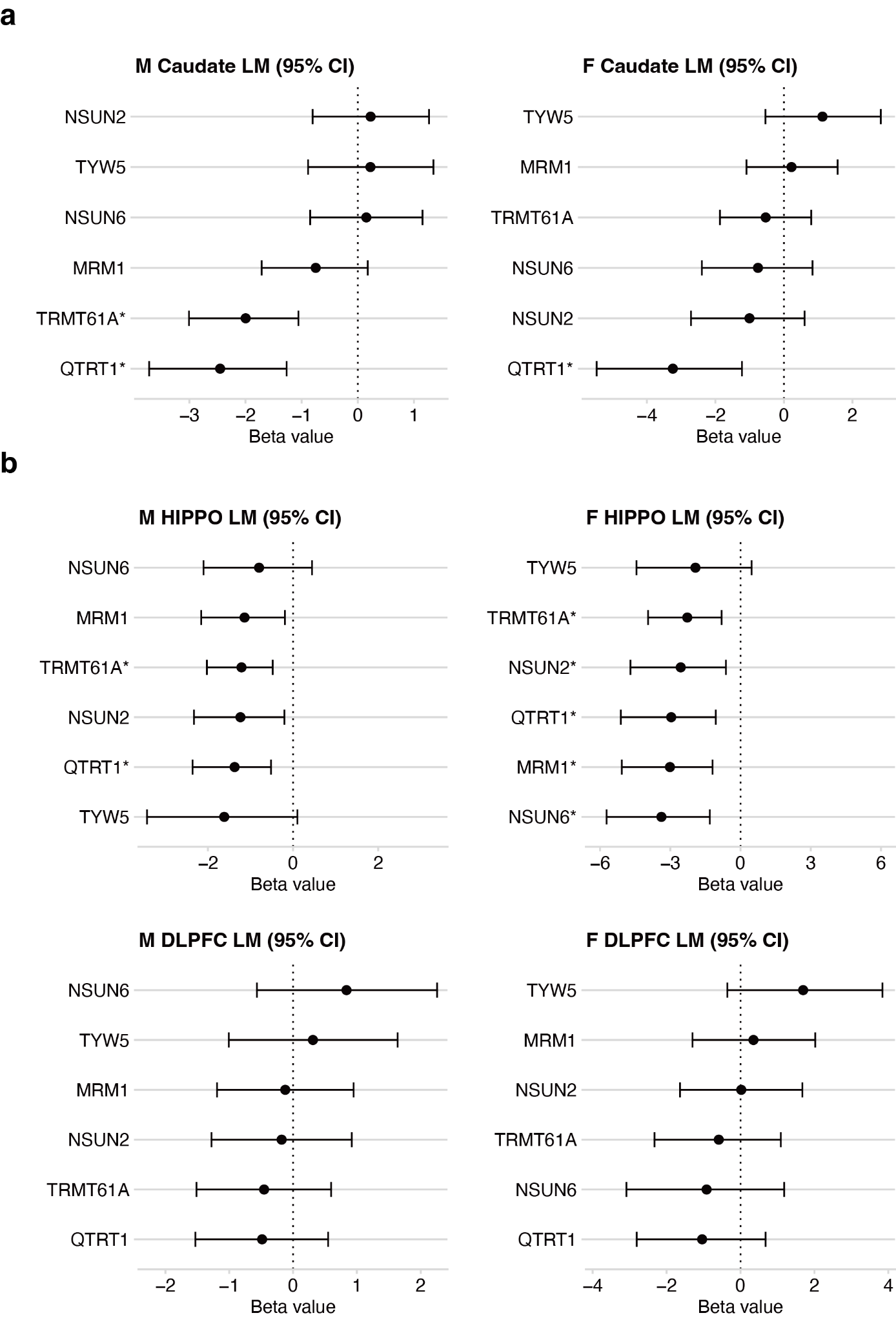

Figure S14: Supplementary figure for Figure 6

Logistic regression analysis of isoform expression-trait associations. Forest plots present the Beta values and 95% confidence intervals (CI) derived from logistic regression models evaluating the association between transcript expression and BIP or SCZ status (*, FDR < 0.05).
